# Metabolomic and Lipidomic Analysis of Manganese-Associated Parkinsonism: a Case-Control Study in Brescia, Italy

**DOI:** 10.1101/2024.09.04.24313002

**Authors:** Freeman Lewis, Daniel Shoieb, Somaiyeh Azmoun, Elena Colicino, Yan Jin, Jinhua Chi, Haiwei Gu, Donatella Placidi, Alessandro Padovani, Andrea Pilotto, Fulvio Pepe, Marinella Turla, Patrizia Crippa, Xuexia Wang, Roberto G Lucchini

**Author notes:** Contributing authors. These authors contributed equally to this work (Freeman Lewis, Daniel Shoieb).

## Abstract

**Background and Objectives:** Excessive Manganese (Mn) exposure is neurotoxic and can cause Mn-Induced Parkinsonism (MnIP), marked by cognitive and motor dysfunction. Although metabolomic and lipidomic research in Parkinsonism (PD) patients exists, it remains limited. This study hypothesizes distinct metabolomic and lipidomic profiles based on exposure status, disease diagnosis, and their interaction.

**Methods:** We used a case-control design with a 2×2 factorial framework to investigate the metabolomic and lipidomic alterations associated with Mn exposure and their link to PD. The study population of 97 individuals was divided into four groups: non-exposed controls (n=23), exposed controls (n=25), non-exposed with PD (n=26) and exposed with PD (n=23). Cases, defined by at least two cardinal PD features (excluding vascular, iatrogenic, and traumatic origins), were recruited from movement disorder clinics in four hospitals in Brescia, Northern Italy. Controls, free from neurological or psychiatric conditions, were selected from the same hospitals. Exposed subjects resided in metallurgic regions (Val Camonica and Bagnolo Mella) for at least 8 continuous years, while non-exposed subjects lived in low-exposure areas around Lake Garda and Brescia city. We conducted untargeted analyses of metabolites and lipids in whole blood samples using ultra-high-performance liquid chromatography (UHPLC) and mass spectrometry (MS), followed by statistical analyses including Principal Component Analysis (PCA), Partial Least Squares–Discriminant Analysis (PLS-DA), and Two-Way Analysis of Covariance (ANCOVA).

**Results:** Metabolomic analysis revealed modulation of alanine, aspartate, and glutamate metabolism (Impact=0.05, p=0.001) associated with disease effect; butanoate metabolism (Impact=0.03, p=0.004) with the exposure effect; and vitamin B6 metabolism (Impact=0.08, p=0.03) with the interaction effect. Differential relative abundances in 3- sulfoxy-L-Tyrosine (β=1.12, FDR p<0.001), glycocholic acid (β=0.48, FDR p=0.03), and palmitelaidic acid (β=0.30, FDR p<0.001) were linked to disease, exposure, and interaction effects, respectively. In the lipidome, ferroptosis (Pathway Lipids=11, FDR p=0.03) associated with the disease effect and sphingolipid signaling (Pathway Lipids=9, FDR p=0.04) associated with the interaction effect were significantly altered. Lipid classes triacylglycerols, ceramides, and phosphatidylethanolamines showed differential relative abundances associated with disease, exposure, and interaction effects, respectively.

**Discussion:** These findings suggest that PD and Mn exposure induce unique metabolomic and lipidomic changes, potentially serving as biomarkers for MnIP and warranting further study.

## 1. INTRODUCTION

In the modern era, characterized by unprecedented industrialization and urban development, the concept of the exposome has emerged as a pivotal framework for understanding the cumulative impact of occupational and environmental exposures on human health [1]. By highlighting the importance of cumulative lifelong exposure, the exposome framework recognizes concomitant factors negatively impacting health outcomes later in life [1]. This perspective has prompted a comprehensive re-evaluation of environmental toxicant mixtures and their additive roles in neurotoxicity development. Among many environmental toxicants, manganese (Mn) has garnered attention [2]. Mn, recognized as an eminent public health concern due to its extensive industrial use and potential for environmental spread, was recently found at unsafe levels in 106 out of 5,034 public water systems (2.1%) in a 2021 Environmental Protection Agency (EPA) survey of North America [3]. In addition, approximately 650 water systems (13%) failed to meet the EPA’s aesthetic standards for taste and appearance, which set a limit of 50 micrograms per liter[3]. These numbers could increase as advancements in steel production and the automotive industry’s shift toward electric vehicles (EVs) drive up demand. Moreover, it should be noted that the EPA’s recommended safety limit is more than twice the levels recommended by national health agencies in Canada (Health Canada and National Institute of Public Health of Quebec (INSPQ)), the World Health Organization, and the European Union [3]. These factors underscore the urgent need for stricter regulatory standards and increased monitoring to mitigate the rising risks associated with Mn exposure [4–6].

The neurotoxic potential of Mn is well-documented in the literature, consistently linking Mn exposure to neurotoxicity and subsequent adverse cognitive and motor health outcomes [7–9]. In alignment with experimental studies, epidemiological studies have demonstrated that communities exposed to chronically elevated levels of Mn exhibit increased risk of Parkinsonian disorders (PDs) [10–12]. These findings underscore Mn’s role as an exogenous trigger for Parkinsonism (PD), particularly in regions with intensive ferroalloy production where cases of manganism have been reported [13]. In response, efforts to identify biomarkers of Mn exposure and effects have commenced, focusing on biofluids and imaging modalities such as magnetic resonance imaging (MRI) [14–16]. However, the search for reliable, reproducible, and easily accessible biomarkers for Mn exposure remains largely unmet. Metabolomics, as an alternative to traditional biomonitoring, is gaining prominence in research literature [17]. In combination with machine learning, metabolomics shows promise for identifying biomarkers of Mn exposure and elucidating the pathways through which Mn exerts toxicity [18–20]. Given the lack of support for many traditional biomarkers, such as Mn levels in blood, plasma, or urine, further exploration of metabolomics is warranted.

Brescia, Italy, a city with historical high industrial activity—especially within the steel and ferroalloy sector—provides a rare opportunity for studying the association between Mn exposure and neurotoxic outcomes. Previous investigations in this region found a higher prevalence of PD among municipalities closer to ferromanganese plants (492/100,000) compared to other province municipalities (321/100,000) [21]. Additionally, a significant association has been found between Bayesian Standardized Morbidity Ratios (SMRs) for PD and dust Mn concentrations [21]. These findings, along with the identification of genetic risk factors linking disruption in Mn intracellular homeostasis to PD, lay the foundation for our study [21]. Building upon this previous work, our study aims to elucidate the metabolomic and lipidomic alterations associated with lifetime Mn exposure and PD. We leverage ultra high-performance liquid chromatography-mass spectrometry (UHPLC-MS) as a research tool to elucidate novel biomarkers of Mn’s neurotoxic effects. We hypothesize that elevated Mn exposure, particularly in historically active industrial regions of Brescia, is associated with distinct changes in metabolomic and lipidomic profiles. The primary outcome of our investigation is the identification of abnormal metabolomic and lipidomic biological processes associated with exposure and/or PD. Our approach not only attempts to bridge the gap in the current understanding of Mn-induced neurotoxicity but also seeks to identify aberrant biological mechanisms that could be targeted with innovative therapeutic and public health interventions.

## 2. MATERIALS AND METHODS

### Study Population

The selection methodology for study participants has been described previously [21]. This study included 97 subjects divided into two groups: 48 exposed subjects (23 cases and 25 controls) and 49 non-exposed subjects (26 cases and 23 controls). Exposed subjects were selected based on having lived outside Brescia (Val Camonica and Bagnolo Mella) for no more than 8 continuous years, considering the half-life of manganese (Mn) in bone [22, 23]. Non-exposed subjects were selected from Lake Garda and Brescia city, non-industrial reference areas within the same province. Exposure levels were determined using GIS to assess proximity to ferromanganese plants, including Mn concentrations in soil, airborne particles, and deposited dust [24]. All subjects underwent fasting whole blood sampling (0.2 ml/sample) at final enrollment. Demographic and lifestyle data, along with clinical diagnosis, treatment, and age at onset for cases, were collected via questionnaires. More detailed study population information can be found in the supplemental material.

### Sample Preparation

Whole blood samples were collected and stored at -80°C by the University of Brescia (UniBS). Following participant consent, Material Transfer and Data Use Agreements compliant with European General Data Protection Regulations, and IRB approval, samples were shared with the FIU Stempel College in 2023. Sample preparation for metabolomics and lipidomics followed previously described methodologies [25–29]. For metabolomics, whole blood samples were thawed at 4°C and 50 µL aliquots were mixed with methanol containing internal standards, incubated at -20C for 20 min, centrifuged, and the supernatant dried and reconstituted for analysis. For lipidomics, blood samples were mixed with PBS, internal standards in methanol, and MTBE, incubated at -20C for 20 min, centrifuged for phase separation, dried, and reconstituted for analysis. More detailed sample preparation information can be found in the supplemental material.

### Untargeted Metabolomic and Lipidomic Data Acquisition

Mass spectrometry experiments were conducted using a Thermo UPLC-Exploris 240 Orbitrap MS (Waltham, MA), with dual injections for both positive and negative ionization modes, and a 1 μL injection volume as described previously [25–29]. Briefly, Metabolomics employed a Waters XBridge BEH Amide column, and lipidomics used a Waters XSelect HSS T3 column, both with specified mobile phases and flow rates. Peak identification involved in-house standards and database searches (HMDB, mzCloud, Metabolika, ChemSpider), with data extraction requiring a minimum absolute intensity of 1,000 and mass accuracy within 5 ppm. Metabolomics data processing used Thermo Compound Discoverer 3.3, while lipidomics used Thermo LipidSearch 4.2 software. More detailed untargeted metabolomic and lipidomic data acquisition information can be found in the supplemental material.

### Statistical Analysis

Descriptive statistics were reported for all covariates, including age, sex, coffee and alcohol consumption, smoking status, and comorbidities. Chi-square tests and t-tests compared groups, while Spearman’s rank correlation analyzed relationships between variables. Metabolomic data analysis included log10-transformation for normalization, Principal Component Analysis (PCA) and Partial Least Squares-Discriminate Analysis (PLS-DA) for exploratory analysis, Permutation Multivariate Analysis of Variance (PERMANOVA) for model evaluation, and Two-Way Analysis of Covariance (ANCOVA) for discerning metabolomic alterations, with False Discovery Rate (FDR) adjustments for p-values. Overrepresentation pathway analysis and metabolite set enrichment analyses were conducted using KEGG and RaMP-DB pathways as suggested by previous reports [30, 31]. Lipidomic data analysis followed similar methods, with biological relevance assessed using LIPEA (https://hyperlipea.org/home) and the KEGG Database, employing Fisher’s exact test and FDR adjustments. Statistical analyses and visualizations were performed using RStudio (Version 2023.09.1+494), MetaboAnalyst (Version 6), and Jupyter Notebooks (Version 6.5.4) accessed through Anaconda. A more detailed statistical analysis plan can be found in the supplemental material.

### Standard Protocol Approvals, Registrations, and Patient Consents

The research plan and consent process received approval from the Ethical Committee of the Civil Hospital in Brescia, Italy. Every participant provided informed consent after receiving both written and oral explanations regarding the study’s objectives and methods. In addition, Institutional Review Board (IRB) approval was obtained through the Office of Research Integrity (ORI) at the Florida International University. The ORI is responsible for the ethical and regulatory oversight of research at Florida International University that involves human subjects. ORI supports and oversees the work of the IRB’s.

### Data Availability

Data not provided in the article because of space limitations may be shared (anonymized) at the request of any qualified investigator for purposes of replicating procedures and results.

## 3. RESULTS

### Demographic and Lifestyle Characteristics of the Study Population

Table 1 compares the characteristics of the study population across two main groups: PD and Controls, each further divided into Exposed and Non-Exposed categories. The Control group included 48 participants (52.1% exposed, 47.9% non-exposed), and the PD group had 49 participants (46.9% exposed, 53.1% non-exposed). Mean ages ranged from 66.4 to 72.4 years, with no significant differences between groups. Males constituted 56.5% to 80.7% of participants, with no significant sex distribution differences. Coffee consumption, alcohol consumption, smoking status, and comorbidities also showed no significant differences, suggesting no association with PD in this population. Geographically, non-exposed participants were from Brescia City and Garda Lake, while exposed participants were predominantly from Valcamonica, with a smaller number from Bagnolo Mella, underscoring a clear distinction between non-exposed and exposed groups. It should be noted that a disproportionate number of participants lived in Valcamonica across both Control and PD groups. Detailed data are available in (*Supplementary Table 1* – *ST1*).

**Table 1:**
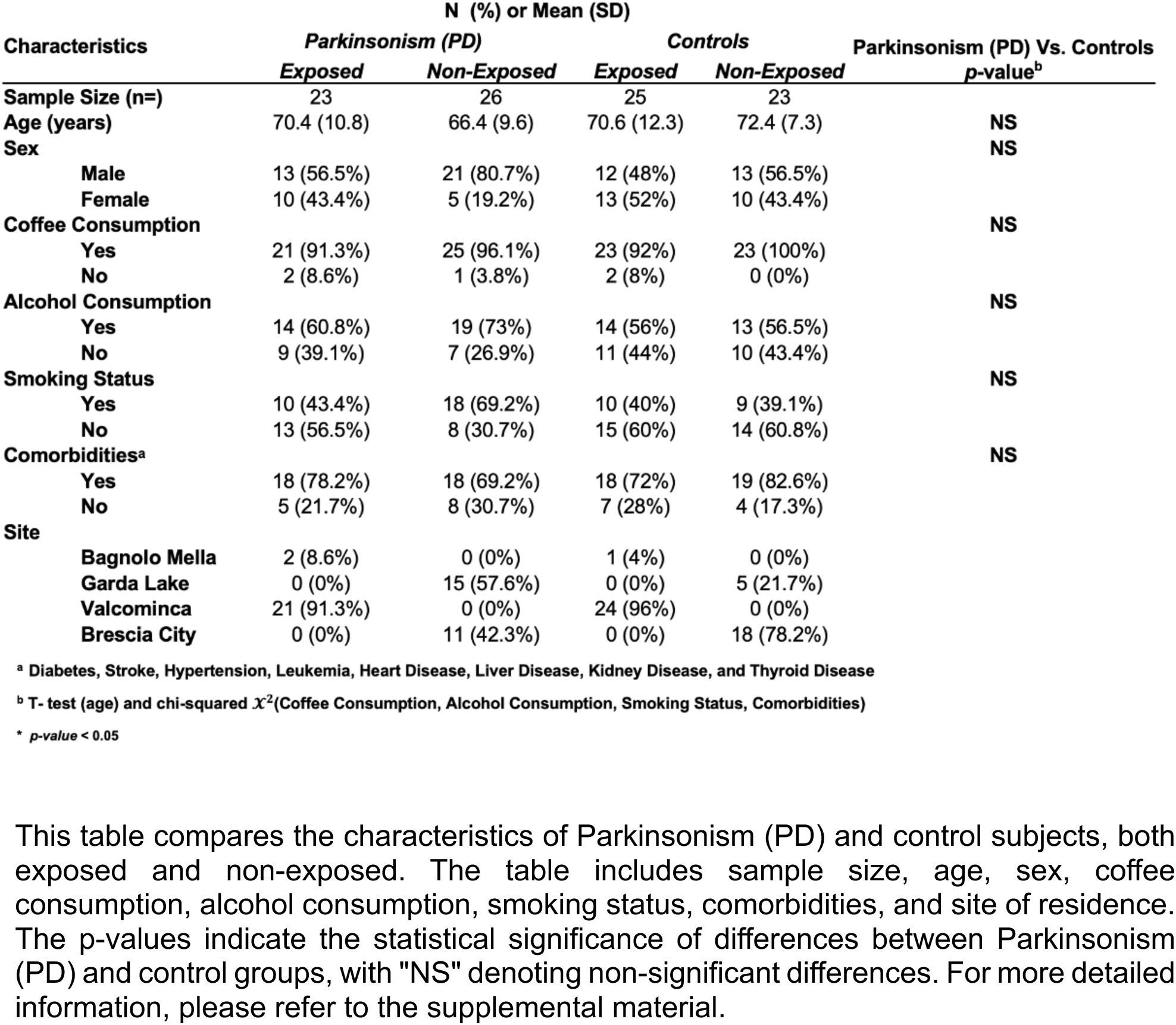
Characteristics of the Study Population.

| Characteristics | N (%) or Mean (SD) |  | N (%) or Mean (SD) |  | Parkinsonism (PD) Vs. Controls<br>p-value<sup>b</sup> |
| --- | --- | --- | --- | --- | --- |
|  | Parkinsonism (PD)<br>Exposed | Parkinsonism (PD)<br>Non-Exposed | Controls<br>Exposed | Controls<br>Non-Exposed |  |
| Sample Size (n=) | 23 | 26 | 25 | 23 |  |
| Age (years) | 70.4 (10.8) | 66.4 (9.6) | 70.6 (12.3) | 72.4 (7.3) | NS |
| Sex |  |  |  |  | NS |
| Male | 13 (56.5%) | 21 (80.7%) | 12 (48%) | 13 (56.5%) |  |
| Female | 10 (43.4%) | 5 (19.2%) | 13 (52%) | 10 (43.4%) |  |
| Coffee Consumption |  |  |  |  | NS |
| Yes | 21 (91.3%) | 25 (96.1%) | 23 (92%) | 23 (100%) |  |
| No | 2 (8.6%) | 1 (3.8%) | 2 (8%) | 0 (0%) |  |
| Alcohol Consumption |  |  |  |  | NS |
| Yes | 14 (60.8%) | 19 (73%) | 14 (56%) | 13 (56.5%) |  |
| No | 9 (39.1%) | 7 (26.9%) | 11 (44%) | 10 (43.4%) |  |
| Smoking Status |  |  |  |  | NS |
| Yes | 10 (43.4%) | 18 (69.2%) | 10 (40%) | 9 (39.1%) |  |
| No | 13 (56.5%) | 8 (30.7%) | 15 (60%) | 14 (60.8%) |  |
| Comorbidities<sup>a</sup> |  |  |  |  | NS |
| Yes | 18 (78.2%) | 18 (69.2%) | 18 (72%) | 19 (82.6%) |  |
| No | 5 (21.7%) | 8 (30.7%) | 7 (28%) | 4 (17.3%) |  |
| Site |  |  |  |  |  |
| Bagnolo Mella | 2 (8.6%) | 0 (0%) | 1 (4%) | 0 (0%) |  |
| Garda Lake | 0 (0%) | 15 (57.6%) | 0 (0%) | 5 (21.7%) |  |
| Valcominca | 21 (91.3%) | 0 (0%) | 24 (96%) | 0 (0%) |  |
| Brescia City | 0 (0%) | 11 (42.3%) | 0 (0%) | 18 (78.2%) |  |

<sup>a</sup> Diabetes, Stroke, Hypertension, Leukemia, Heart Disease, Liver Disease, Kidney Disease, and Thyroid Disease
<sup>b</sup> T- test (age) and chi-squared $\chi^2$ (Coffee Consumption, Alcohol Consumption, Smoking Status, Comorbidities)
\* p-value < 0.05
This table compares the characteristics of Parkinsonism (PD) and control subjects, both exposed and non-exposed. The table includes sample size, age, sex, coffee consumption, alcohol consumption, smoking status, comorbidities, and site of residence. The p-values indicate the statistical significance of differences between Parkinsonism (PD) and control groups, with "NS" denoting non-significant differences. For more detailed information, please refer to the supplemental material.

### Metabolomics

Analysis of Covariance (ANCOVA) and Metabolite Associations with Disease Effect Following exploratory analysis, ANCOVA identified 156 metabolites significantly associated with the disease, reduced to 99 after FDR correction (*Supplementary Figure 6 – SF6*). This indicates that even after stringent statistical correction, many metabolites display changes that are potentially characteristic of the disease state. The volcano plot (*Figure 1A*) contrasts the significance of observed changes in relative abundance (indicated by p < 0.05) with their effect size (represented by beta coefficients), for the disease effect. Notably, metabolites such as “dimethyl sulfoxide” (β=-1.31, FDR p<0.001), “sulbactam” (β=1.61, FDR p<0.001), and “dopamine 3-O-sulfate” (β=1.15, FDR p<0.001) exhibited high statistical significance, suggesting a strong association with the disease.

**Figure 1:**
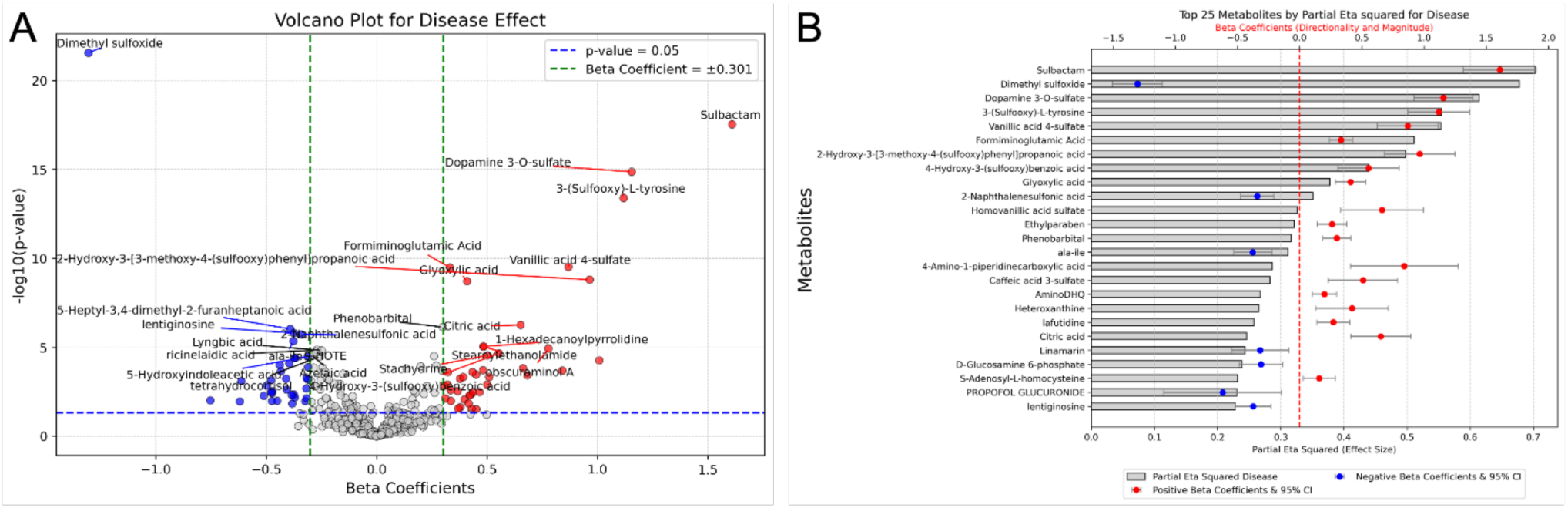
Disease Effect on Metabolites. Panel A: Volcano plot for disease effect, showing the relationship between the beta coefficients (effect size) and the -log10(p-value) of metabolites. Red points represent metabolites with significant positive associations, and blue points represent significant negative associations with the disease effect. The vertical green line indicates a beta coefficient of ± 0.301, and the horizontal blue line indicates a significance threshold of p = 0.05. Panel B: Bar chart of the top 25 metabolites significantly associated with the disease effect, ranked by their partial eta squared 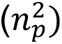 values. The bars represent the magnitude of effect, with red points indicating parameter estimates (beta coefficients) and error bars showing the 95% confidence intervals. The table includes metabolites’ names and their corresponding effect sizes.

The labeled metabolites were limited to only the top 25 metabolites statistically significant after FDR adjustment for legible reporting (a table with the full list of metabolites is in Supplementary Table 2 – ST2). The bar chart (*Figure 1B*) represents the top 25 metabolites associated with the disease hierarchically ranked by their 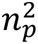 values. Again, “sulbactam” 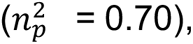 “dimethyl sulfoxide” 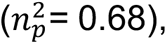 and “dopamine 3-O-sulfate” 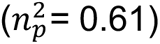 demonstrate the largest magnitude of effect associated with disease (a table with the full list of 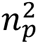 values can be found in *ST2*). The pathway analysis bubble plot (*Supplementary Figure 8 – SF8*) employs a bubble plot visualization to evaluate the significance and impact of metabolic pathways associated with the disease effect. Notably, “alanine, aspartate and glutamate metabolism” (Impact=0.05, p=0.001), the “citrate cycle (TCA cycle)” (Impact=0.15, p=0.01), “glyoxylate and dicarboxylate metabolism,” (Impact=0.31, p=0.03), and “arginine and proline metabolism” (Impact=0.02, p=0.03) exhibited statistically significant association with the disease effect (a table with the full list of pathways can be found in *Supplementary Table 3 – ST3*). *(SF8)* also presents a bubble plot that depicts the results of the MSEA using the RaMP-DB pathway library. The bubble plot highlights enriched pathways related to amino acid metabolism, such as “Alanine, aspartate and glutamate metabolism” (Expect=0.352, p=0.006) and energy production, such as “Citric acid cycle and respiratory electron transport” (Expect=0.255, p=0.002) (a table with the full list of MSEA pathways can be found in *Supplementary Table 4 – ST4)*. These biological processes, markedly implicated in the main effect of disease, may contribute to the etiology and pathogenesis of PD, representing critical areas for potential further study

### ANCOVA and Metabolite Associations with Exposure Effect

We identified 29 statistically significant metabolites related to exposure after FDR correction (*SF6*). As demonstrated in the volcano plot (*Figure 2A*), “dimethyl sulfoxide” (β=-0.91, FDR p<0.001), “D-(-)-Mannitol” (β=0.64, FDR p<0.001), and “L-Iditol” (β=0.86, FDR p<0.001), emerged as notable metabolites with a substantial association with the exposure effect. Additionally, “L-Histidinol phosphate” (β=0.59, FDR p=0.002), “lyngbic acid” (β=-0.24, FDR p=0.002), and “citric acid” (β=0.51, FDR p=0.003), demonstrated a substantial statistical significance suggesting a marked association with exposure. *(Supplementary Figure 9 - SF9*) illustrates the top 25 metabolites ranked by their 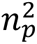 values, illustrating magnitude of effect of exposure on each metabolite. “Dimethyl sulfoxide” 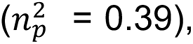 “tetrahomomethionine” 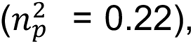 and “lafutidine” 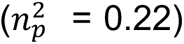 exhibit higher 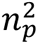 values, suggesting the strongest effect of exposure on their relative abundance. The pathway analysis bubble plot (*Figure 2B*) reveled “alanine, aspartate and glutamate metabolism” (Impact=0.2, p<0.001), “butanoate metabolism” (Impact=0.03, p=0.004), and “glyoxylate and dicarboxylate metabolism” (Impact=0.03, p=0.01) as statistically significant, indicating the largest effect of exposure on these pathways. *(SF9*) also represents the top 25 metabolite sets enriched due to exposure, as determined by MSEA. The results of the enrichment analysis revealed several metabolites sets with significant alterations related to transmembrane transporters. “SLC-mediated transmembrane transport” (Expect=0.326, p=0.003), “transport of small molecules” (Expect=0.437, p=0.007), and “sodium-coupled sulphate, di- and tri-carboxylate transporters” (Expect=0.00841, p=0.008) showed higher levels of enrichment, indicating a significant perturbation in these metabolite sets due to exposure. Note, biological relevance should be interpreted with caution as no pathway or enriched set achieved statistical significance after FDR correction. All corresponding primary exposure effect data can be found in (*ST2 – ST4*).

**Figure 2:**
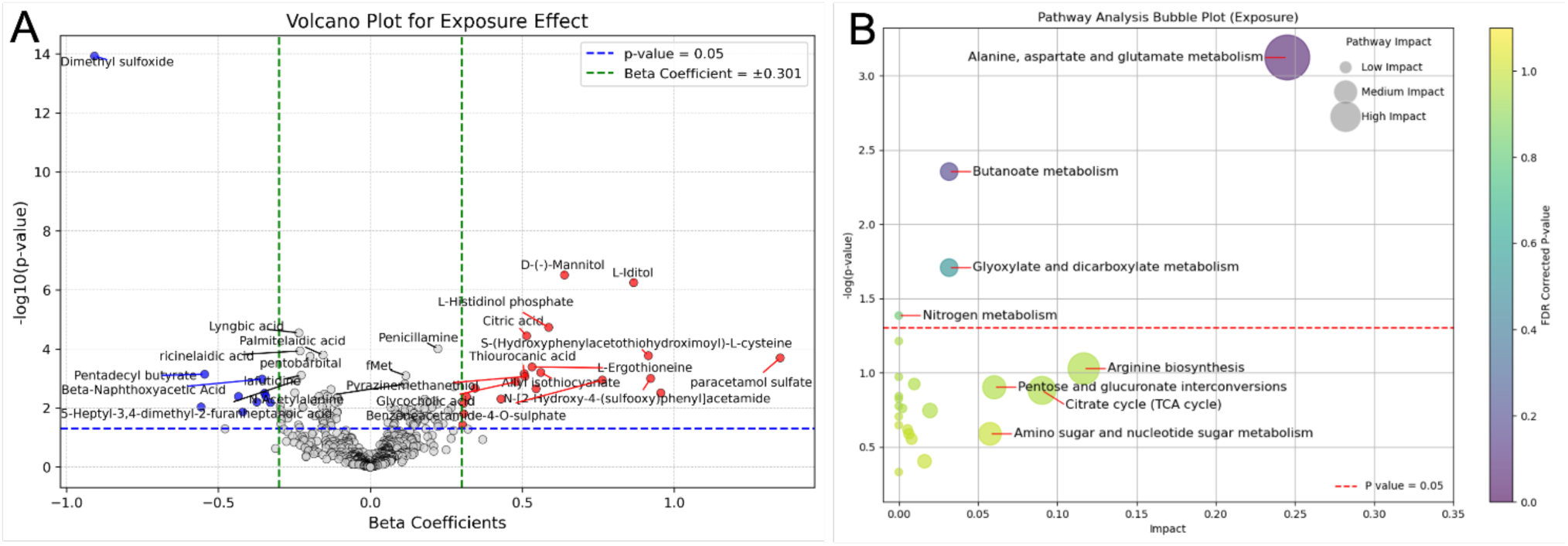
Exposure Effect on Metabolites. Panel A: Volcano plot for the exposure effect, showing the relationship between the beta coefficients (effect size) and the -log10(p-value) of metabolites. Red points represent metabolites with significant positive associations, and blue points represent significant negative associations with the exposure effect. The vertical green line indicates a beta coefficient of ± 0.301, and the horizontal blue line indicates a significance threshold of p = 0.05. Panel B: Pathway analysis bubble plot for exposure effect, highlighting the significance and impact of various metabolic pathways. Pathways with high impact and significance include “Alanine, aspartate and glutamate metabolism” and “Glyoxylate and dicarboxylate metabolism.” The color gradient represents the FDR corrected p-value, with pathways above the red dashed line (p-value = 0.05) considered statistically significant.

### ANCOVA and Metabolite Associations with Interaction Effect

We found 58 statistically significant metabolites associated with the interaction effect between disease and exposure after FDR correction (SF6). The volcano plot (*Figure 3A*) highlights key metabolites such as “palmitelaidic acid” (β=0.30, FDR p<0.001), “pentobarbital” (β=0.35, FDR p<0.001), “(S)-2-methlbutanal (β=0.60, FDR p<0.001), “dimethyl sulfoxide” (β=0.68, FDR p<0.001), “lyngbic acid” (β=0.36, FDR p<0.001) and “n-Butyl lactate” (β=0.60, FDR p<0.001), showing a positive association with the interaction effect. Markedly, many of the statistically significant metabolites associated with the interaction effect showed a positive association. Notably, “dimethyl sulfoxide” reversed its association from negative in main effects to positive in the interaction context. In *(Supplementary Figure 10 – SF10) “*palmitelaidic acid” 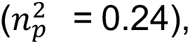 “(S)-2-methylbutanal” 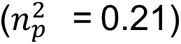 and “n-Butyl lactate” 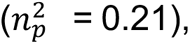 displayed the largest 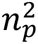 values among the metabolites, indicating that a substantial portion of their variance is explained by the interaction effect. Pathway analysis bubble plot (*SF10*) revealed “vitamin B6 metabolism” (Impact=0.08, p=0.03) as significant, although not after FDR correction. Additionally, “tyrosine metabolism” (Impact=0.13, p=0.15) is associated with the interaction effect; however, the impact does not reach the level of significance. It should be noted that a considerably lower number of pathways were identified compared to the main effects of disease (*SF8*) and exposure (*Figure 2B)*. MSEA (*Figure 3B*) broadly identified amino acid metabolism and transmembrane transporters as enriched metabolite sets. These findings corroborate what was previously found in our main effect of disease *(SF8*) and main effect of exposure (*SF9*). “phase II - Conjugation of compounds” (Expect=0.341, p<0.001), “sudden infant death syndrome (SIDS) susceptibility pathways” (Expect=0.0327, p<0.001), and “SLC transporter disorders” (Expect=0.187, p<0.001) were among the top three statistically significant enriched sets. However, the smaller size of their bubbles indicates a low enrichment ratio. Conversely, high enrichment ratios were found among “defective SLC22A12 causes renal hypouricemia 1 (RHUC1)” (Expect=0.00467, p=0.005), “defective SLC6A3 causes Parkinsonism-dystonia infantile (PKDYS)” (Expect=0.00467, p=0.005), and “defective SLC35A1 causes congenital disorder of glycosylation 2F (CDG2F)” (Expect=0.00467, p=0.005). Of interest is the “defective SLC6A3 causes Parkinsonism-dystonia infantile (PKDYS)” enriched set due to its relevance to PD. All corresponding primary interaction effect data can be found in (*ST2 – ST4*).

**Figure 3:**
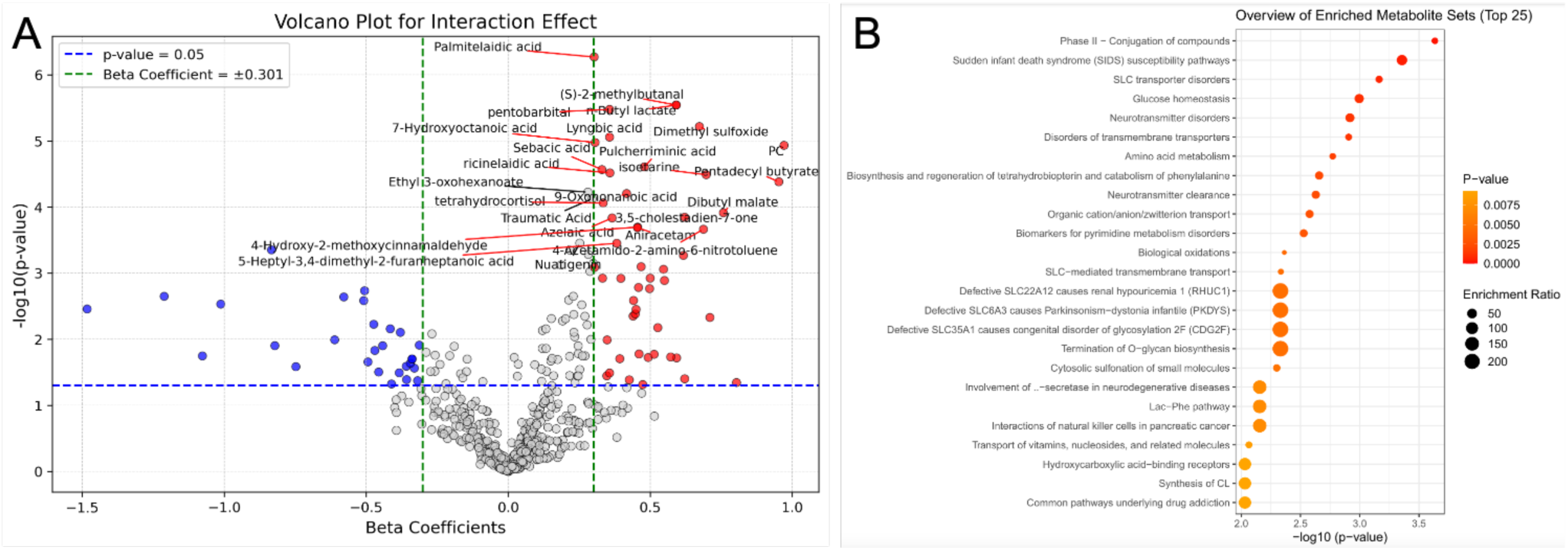
Interaction Effect on Metabolites. Panel A: Volcano plot for the interaction effect, showing the relationship between the beta coefficients (effect size) and the -log10(p-value) of metabolites. Red points represent metabolites with significant positive associations, and blue points represent significant negative associations with the interaction effect. The vertical green line indicates a beta coefficient of ± 0.301, and the horizontal blue line indicates a significance threshold of p = 0.05. Panel B: Bubble plot of the top 25 enriched metabolite sets for the interaction effect. The size of the bubbles represents the enrichment ratio, and the color gradient indicates the p-value. More significant pathways are highlighted with larger and darker- colored bubbles.

### Lipidomics

#### ANCOVA and Lipid Associations with Disease Effect, Exposure Effect, and Interaction Effect

Transitioning from our metabolomic analysis, our ANCOVA identified 33 lipids significantly associated with the disease effect after FDR correction (*Supplementary Figure 13 – SF13*). The volcano plot (*Figure 4A*) shows notable lipids such as “TG(16:0_10:0_18:1)” (β=0.8, FDR p=0.001), “SM(d44:3)” (β=0.17, FDR p=0.01), and “TG(18:0_18:0_18:1)” (β=0.37, FDR p=0.01) with positive associations (a table with the full list of lipids is in *Supplementary Table 5 – ST5*). The bar chart (*Figure 4B*) ranks the top 25 lipids by 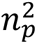 values, highlighting “TG(16:0_10:0_18:1)” 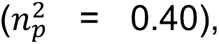 “TG(16:1_14:0_18:2)” 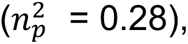 and “TG(16:0_14:0_16:0)” 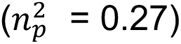 as having the largest effects (a table with the full list of 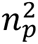 values can be found in *ST5*). For the exposure effect, no statistically significant lipids were identified after FDR correction, but notable lipids included “Cer(d18:1_24:0)” (β=-0.87, FDR p=0.25) and “Cer(d18:0_24:0)” (β=-0.12, FDR p=0.25) (*Figure 4C*). The bar chart (*Figure 4D*) ranks the top 25 lipids by 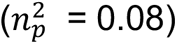 values, with “Cer(d18:1_24:1)” 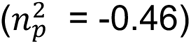 showing the largest effect. The interaction effect revealed 12 significant lipids after FDR correction (*SF13*), including “PE(16:0_20:4)” (β=-0.39, FDR p=0.02) and “PE(40:7e)” (β=-0.38, FDR p=0.02) (*Figure 4E*). The bar chart (Figure 4F) ranks the top 25 lipids by 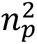 values, with “PC(37:5e)” 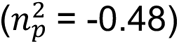 and “PE(40:5e)” 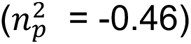 having the largest negative associations. Pathway analysis (*Supplementary Table 6* – *ST6*) linked the disease effect to retrograde endocannabinoid signaling (Pathway Lipids=8 p=0.03), ferroptosis (Pathway Lipids=11, p=0.03), and glycerophospholipid metabolism (Pathway Lipids=26, p=0.05). The interaction effect was associated with retrograde endocannabinoid signaling (Pathway Lipids=8, p=0.04), ferroptosis (Pathway Lipids=11, p=0.04), and sphingolipid signaling pathway (Pathway Lipids=9, p=0.04). The exposure effect was not included due to the absence of significant lipids. All corresponding primary data can be found in *ST5* and *ST6*.

**Figure 4:**
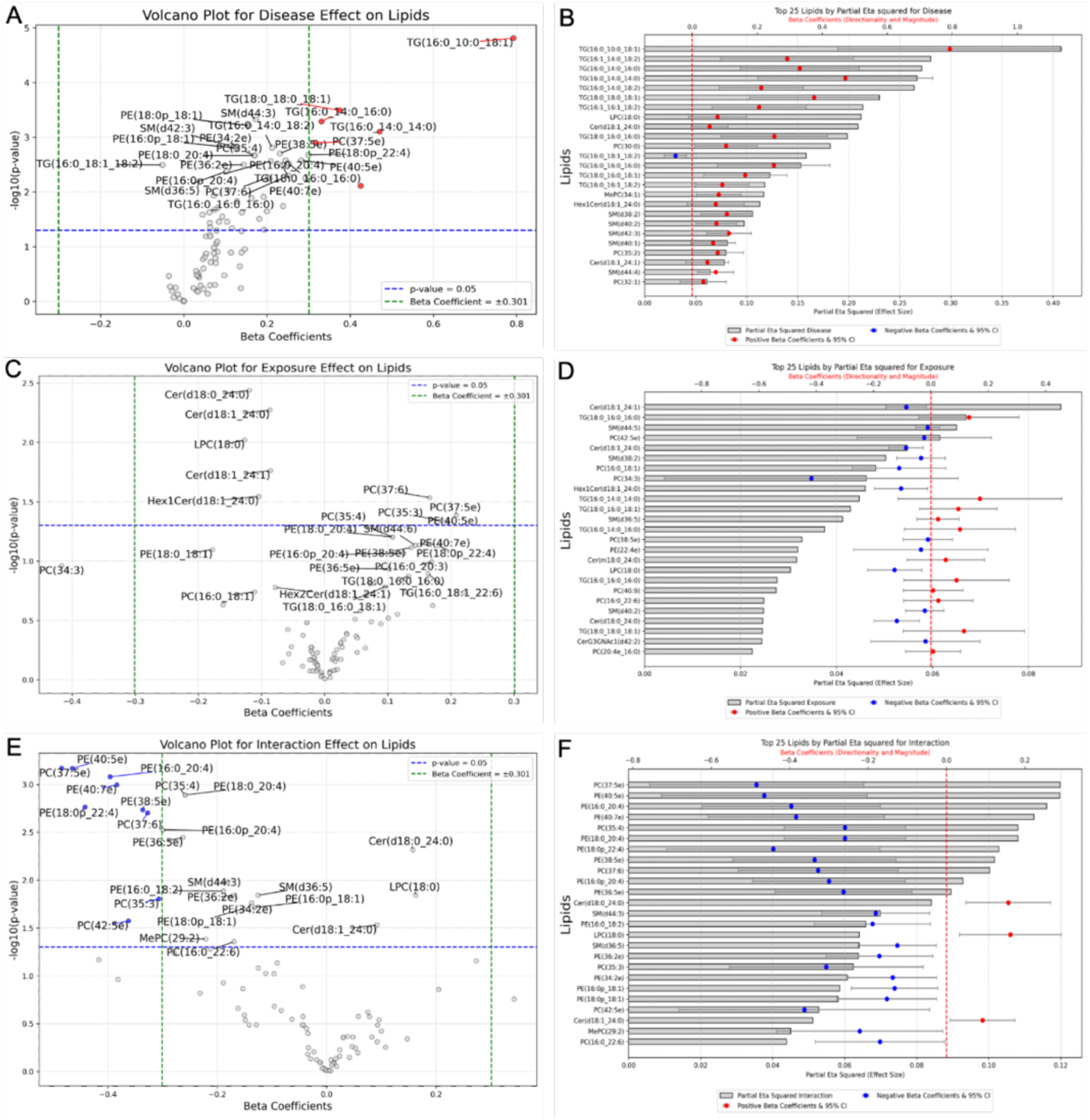
Volcano Plots and Bar Charts for Lipid Effects. Panel A: Volcano plot for disease effect on lipids, showing the relationship between the beta coefficients (effect size) and the -log10(p-value) of lipids. Significant positive associations are marked in red, and negative associations are marked in blue. The vertical green line indicates a beta coefficient of ± 0.301, and the horizontal blue line indicates a significance threshold of p = 0.05. Panel B: Bar chart of the top 25 lipids significantly associated with the disease effect, ranked by their partial eta squared 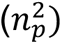 values. Red points indicate parameter estimates (beta coefficients), and error bars show the 95% confidence intervals. Panel C: Volcano plot for exposure effect on lipids, like Panel A, highlighting significant associations with the exposure effect. Panel D: Bar chart of the top 25 lipids significantly associated with the exposure effect, with rankings and visual representation like Panel B. Panel E: Volcano plot for interaction effect on lipids, showing the combined impact of disease and exposure on lipid levels. Panel F: Bar chart of the top 25 lipids significantly associated with the interaction effect, presented similarly to Panels B and D.

## 4. DISCUSSION

These findings suggest that alterations in metabolite and lipid signatures could serve as predictive biomarkers for specific PD subtypes related to environmental exposure. These insights into the metabolomic and lipidomics perturbations associated with PD and environmental exposure may aid in preventive screenings, novel diagnostic methods, and therapeutic strategies. While statistical significance is essential, biological relevance is equally important in identifying clinically relevant biomarkers. Thus, we focus our discussion on metabolomic and lipidomic perturbations that are both statistically significant and biologically relevant, as represented in (*Table 2*).

**Table 2:** Summary of Significant Metabolites and Pathways for Disease, Exposure, and Interaction Effects.

|  | DISEASE EFFECT | EXPOSURE EFFECT | INTERACTION EFFECT |
| --- | --- | --- | --- |
| Metabolomics | 3-sulfoxy-L-Tyrosine | glycocholic acid | palmitelaidic acid |
|  | formiminoglutamic acid | butanoate | DMSO |
|  | glyoxylic acid | glutamate | vitamin B6 metabolism |
|  | DMSO | DMSO | glucose homeostasis |
|  | amino acid metabolism | butanoate metabolism | amino acid metabolism |
|  | TCA cycle | alanine, aspartate, and glutamate metabolism | SLC transporters disorders |
|  |  | SLC-mediated transmembrane transport |  |
| Lipidomics | triacylglycerols | ceramides | phosphatidylethanolamines |
|  | Ferroptosis |  | ferroptosis |
|  | Endocannabinoid |  | endocannabinoid |
|  |  |  | sphingolipid metabolism and signaling |

This table provides an overview of the significant metabolites and pathways associated with the disease, exposure, and interaction effects. The rows categorize the findings into metabolomics and lipidomics, while the columns delineate the specific effects.

### Disease Effect on Metabolites

Significant metabolites related to medication usage, dietary habits, amino acid metabolism, cellular redox balance, and vitamin B regulation were identified. However, some metabolites like sulbactam, dopamine 3-O-sulfate (DA-3S), vanillic acid, and 3- hydroxy-3-[3-methoxy-4-(sulfoxy)-phenyl] propanoic acid (3HMSPP) may be biologically artifactual. For example, sulbactam, a beta-lactamase inhibitor, is often used by PD patients to treat infections [32–34] and DA-3S, a dopamine metabolite, reflects increased dopamine metabolism but shows no consistent link with PD severity [35, 36]. Similarly, vanillic acid and 3HMSPP, found in common foods and beverages, are not likely to serve as diagnostic biomarkers [37, 38]. Conversely, biologically relevant metabolites such as 3-sulfoxy-L-tyrosine, formiminoglutamic acid, glyoxylic acid, and dimethyl sulfoxide (DMSO) were identified. 3-sulfoxy-L-tyrosine, a post-translational modification of tyrosine, plays a role in norepinephrine synthesis, which may be reduced in PD [39, 40].

Formiminoglutamic acid, an intermediate in L-histidine breakdown, serves as a biomarker for folate levels, and its increase can indicate vitamin B12 deficiency, which is associated with PD [41]. Glyoxylic acid, crucial for cellular redox balance and amino acid metabolism, is linked to mitochondrial function and neuroprotection, highlighting its importance in PD [42]. Lastly, DMSO, known for its anti-inflammatory and antioxidant properties, might play a protective role in neurodegenerative processes, though its exact impact in PD requires further investigation [43, 44]. The disease effect also impacted pathways related to amino acid metabolism and the TCA cycle. Alterations in 3-sulfoxy-L-tyrosine, formiminoglutamic acid, and glyoxylic acid suggest that nerve degeneration and mitochondrial dysfunction could lead to changes in amino acid metabolism, exacerbating PD symptoms [45, 46]. These findings align with existing literature and underscore the complex interplay between neurodegeneration and metabolic dysfunction.

### Exposure Effect on Metabolites

Tetrahomomethionine, lafutidine, and mannitol were strongly associated with the exposure effect but were excluded as diagnostic or prognostic biomarkers due to their artifactual nature and lack of biological relevance. Lafutidine and mannitol are strongly correlated with medication usage, while tetrahomomethionine is linked to dietary intake of L-methionine, though its biosynthetic pathway is unclear [47, 48]. Glycocholic acid, a bile acid conjugated with glycine, is upregulated, possibly due to liver damage caused by metal exposure, which aligns with its role in emulsifying fats and enhancing solubility [49, 50]. Butanoate metabolism was significantly altered, reflecting the breakdown of L-glutamate into GABA. Metals, particularly manganese (Mn), disrupt these pathways by interfering with neurotransmitter systems, including blocking NMDA calcium channels and inhibiting the expression of NMDAR subunits (GluN1, GluN2A, GluN2B). Mn exposure also induces alpha-synuclein overexpression, leading to phosphorylation and downregulation of the GluN2B subunit, impairing NMDAR signaling. Additionally, Mn decreases GABAA receptor expression and induces GABAB receptors, contributing to alpha-synuclein accumulation [49, 51]. Metal exposure also disrupts SLC-mediated transport, activating proinflammatory genes (e.g., IL-6, IL-1B, CCL2), competing with essential metal ions, and blocking TRPC3 channels in astrocytes. Moreover, metals alter amino acid metabolism by binding to sites, catalyzing oxidation, disrupting protein folding, and displacing essential ions. These disruptions likely drive the alterations in amino acid metabolism observed with the exposure effect [49, 52]. Furthermore, higher doses of metals directly damage intestinal flora by causing cell death through homeostasis imbalances and intracellular interactions with vital proteins and DNA [53]. Alterations in oxidative stress signaling and downregulation of DMSO (related to thiol-disulfide homeostasis) were also noted. Metal exposure increases free radicals via Fenton & Haber-Weiss reactions, where metal ions catalyze the conversion of hydrogen peroxide into hydroxyl radicals, elevating oxidative stress. Glutathione, a key intracellular antioxidant, is disrupted, further affecting redox balance. The downregulation of DMSO, formed from dimethyl sulfide reacting with reactive oxygen species (ROS), may heighten oxidative stress [54]. Lastly, exposure effects on taste perception were observed. Phantogeusia, a taste disorder, is a recognized symptom in areas affected by heavy metal fume exposure [55].

### Interaction Effect on Metabolites

N-butyl lactate, 2-methylbutanal, and pentobarbital were statistically associated with the interaction effect between disease and exposure status but were considered artifactual due to confounding factors such as diet and medication [56, 57]. However, palmitelaidic acid and DMSO were both statistically significant and biologically relevant. Palmitelaidic acid, a trans fatty acid associated with increased cardiovascular disease risk, is primarily obtained through diet. It also plays a role in lipid peroxidation, a process where ROS cause oxidative degradation of lipids, leading to oxidative stress, cell damage, and PD disease progression [58, 59]. Key pathways affected by the interaction effect—glucose homeostasis, amino acid metabolism, and SLC transporter disorders— highlight how combined exposure and disease effects mirror biological perturbations seen in the independent main effects, presumably heightening them. Uniquely, vitamin B6 metabolism was significantly impacted, highlighting its importance in amino acid metabolism and neurotransmitter synthesis, which are crucial for proper brain function [60]. Additionally, impairments in glucose homeostasis, supported by previous PD patient literature, was associated with the interaction effect, potentially due to mechanisms like insulin resistance, oxidative stress, and blood-brain barrier dysfunction [61]. In summary, palmitelaidic acid and DMSO, which have been previously linked to both disease and exposure effects, and are also associated with the interaction effect, may serve as potential diagnostic or prognostic biomarkers warranting further investigation.

### Disease and Interaction effect on Lipids

Lipid classes were significantly altered in association with the disease and interaction effects, while the exposure effect did not reach statistical significance. This suggests a greater lipid class alteration when disease and exposure effects are combined. Key lipids involved include triglycerides (TG), lysophosphatidylcholine (LPC), ceramides, and phosphatidylcholines (PC), all of which are linked to cognitive function [62]. Longitudinal studies indicate that elevated triglyceride levels increase the risk of cognitive impairment, a hallmark of late-stage PD [63]. Ceramides, a class of sphingolipids (SPs), are crucial for cellular processes like division, differentiation, and apoptosis. Dysregulation in sphingolipid metabolism is associated with neurological disorders and disease progression [64, 65]. LPC, a phospholipid integral to cell membrane structure and function, plays roles in signaling, inflammation, and immune regulation. Abnormal LPC levels have been linked to neurological disorders and may reflect oxidative stress response [66]. The interaction effect interestingly showed a downregulation in lipid classes, especially phosphatidylethanolamine (PE), corroborating findings in other PD studies [67]. These altered lipids are associated with pathways like retrograde endocannabinoid signaling, ferroptosis, and sphingolipid metabolism [68–70]. The disease and interaction effects significantly impacted endocannabinoid signaling and ferroptosis, with the interaction effect also affecting sphingolipid metabolism. Endocannabinoids, such as anandamide (AEA) and 2-arachidonoylglycerol (2AG), regulate synaptic activity and mitochondrial function through CB1 receptors [68]. Ferroptosis, a form of regulated cell death driven by ROS and lipid peroxidation, is involved in various pathological processes, including PD [69]. Lastly, sphingolipids, like ceramide and sphingosine-1-phosphate (S1P), play opposing roles in cell stress responses and survival, highlighting their importance in these pathways [70].

This study provides valuable insights into the metabolomic and lipidomic alterations associated with manganese exposure and PD. Our findings highlight distinct metabolic and lipid signatures, including the potential roles of palmitelaidic acid and DMSO as diagnostic or prognostic biomarkers, which could significantly contribute to the early detection and management of manganese-induced Parkinsonism (MnIP). The study’s strengths lie in its comprehensive approach, using a case-control design and advanced metabolomic and lipidomic analyses, which allow for the identification of potential biomarkers associated with both disease and exposure effects. However, this study has limitations that should be acknowledged. A significant limitation is the cross-sectional nature of the study and the fact that the participants were already diagnosed with PD, which precludes establishing causality between manganese exposure and disease onset. Additionally, the relatively small sample size may limit the generalizability of the findings. Future studies should focus on larger, longitudinal cohorts to better understand the temporal relationship between exposure and disease development and to validate the identified biomarkers. The current findings will be instrumental in conducting a power analysis to determine the sample size needed for future studies, ensuring that they are adequately powered to detect meaningful differences and associations. Future research should also incorporate transcriptomic and proteomic analyses to further investigate the metabolic and lipid pathways identified in this study. Integrating these omics approaches will enable a more comprehensive understanding of how manganese exposure disrupts metabolic homeostasis and contributes to neurodegenerative processes. In terms of public health significance, our study underscores the urgent need for stricter regulatory standards and enhanced monitoring of environmental manganese exposure, particularly in industrial regions. The identification of biomarkers like palmitelaidic acid and DMSO could pave the way for improved screening protocols and targeted interventions aimed at mitigating the neurotoxic effects of manganese. Furthermore, understanding the metabolic pathways disrupted by manganese exposure may open new avenues for therapeutic strategies to prevent or slow the progression of PD in affected populations.

## Author Contributions

Freeman Lewis, MPH

Environmental Health Sciences, Florida International University, Miami, Florida, USA

Drafting/revision of the manuscript, including medical writing for content; study concept or design; analysis and interpretation of data

Daniel Shoieb, MD

Department of Medical and Surgical Specialties, Radiological Sciences and Public Health, University of Brescia, Brescia, Italy

Drafting/revision of the manuscript, including medical writing for content; study concept or design; and interpretation of data

Somaiyeh Azmoun, PhD

Environmental Health Sciences, Florida International University, Miami, Florida, USA

Revision of the manuscript; data acquisition

Elena Colicino, PhD

Department of Environmental Medicine and Climate Science, Icahn School of Medicine at Mount Sinai, New York, New York, USA

Revision of the manuscript for content, including medical writing for content; study concept or design; interpretation of analysis; supervision

Yan Jin, PhD

Environmental Health Sciences, Florida International University, Miami, Florida, USA

Revision of the manuscript for content, data acquisition; and interpretation of data

Jinhua Chi, PhD

Environmental Health Sciences, Florida International University, Miami, Florida, USA

Data acquisition and design

Haiwei Gu, PhD

Environmental Health Sciences, Florida International University, Miami, Florida, USA

Revision of the manuscript for content, including medical writing for content; major role in the acquisition of data; interpretation of data; resources; supervision

Donatella Placidi, PhD

Department of Medical and Surgical Specialties, Radiological Sciences and Public Health, University of Brescia, Brescia, Italy

Revision of the manuscript for content, including medical writing for content; major role in the acquisition of data; study concept or design; interpretation of data; resources; supervision

Alessandro Padovani, MD, PhD

Department of Clinical and Experimental Sciences, University of Brescia, Brescia, Italy; Department of Continuity of Care and Frailty, Neurology Unit, ASST Spedali Civili Hospital, Brescia, Italy

Revision of the manuscript for content, including medical writing for content; study concept or design; data acquisition; resources

Andrea Pilotto, MD

Department of Clinical and Experimental Sciences, University of Brescia, Brescia, Italy; Department of Continuity of Care and Frailty, Neurology Unit, ASST Spedali Civili Hospital, Brescia, Italy

Revision of the manuscript for content, including medical writing for content; major role in the acquisition of data; study concept or design; interpretation of data; resources; supervision

Fulvio Pepe, MD

Clinic of Neurology, Poliambulanza Foundation, Brescia, Italy Major role in the acquisition of data; and resources

Marinella Turla, MD

Clinic of Neurology, Esine Hospital of Valcamonica, Brescia, Italy

Major role in the acquisition of data; and resources

Xuexia Wang, PhD

Department of Biostatistics, Florida International University, Miami, Florida, USA

Revision of the manuscript for content; and interpretation of data

Roberto G. Lucchini, MD

Environmental Health Sciences, Florida International University, Miami, Florida, USA; Department of Biomedical, Metabolic and Neurosciences, University of Modena and Reggio Emilia, Modena, Italy

Revision of the manuscript for content, including medical writing for content; study concept or design; major role in the acquisition of data; resources; and supervision

