## Supplemental Material for "Metabolomic and Lipidomic Analysis of Manganese-Associated Parkinsonism: a Case-Control Study in Brescia, Italy"

Supplemental Material and Methods

### Study Population

Study participant selection methodology has been described previously [21]. For this study, population includes 97 subjects divided into two groups: 48 exposed subjects (23 cases and 25 controls) and 49 non-exposed subjects (26 cases and 23 controls). The exposure level was determined based on the proximity to ferromanganese plants using Geographical Information Systems (GIS). This assessment included detailed spatial analysis to measure the distance of subjects' residences from the ferromanganese plants. Additionally, environmental measures were considered, including concentrations of Mn in soil samples, levels of airborne particles, and deposited dust in the areas surrounding the plants[23]. This selection criteria provided a comprehensive evaluation of the environmental exposure to Mn for each subject. Exposed subjects were selected based on having lived outside Brescia (Val Camonica and Bagnolo Mella) for no more than 8 continuous years of their lifetime, considering the half-life of Mn stored in bone [22, 23]. Non-exposed subjects were selected from Lake Garda and Brescia city, which are non-industrial reference areas within the same province of Brescia. All selected subjects underwent fasting whole blood sampling (0.2 ml/sample) for blood biomarker analyses at final enrollment. Self-report data through questionnaire on demographics (age and sex), lifestyle habits (coffee consumption, alcohol consumption, smoking status, and prior diagnosis of comorbid diseases), and, exclusively to the cases, clinical diagnosis, treatment, and age at onset was collected.

### Sample Preparation

Whole blood samples were collected at final enrollment and stored at -80˚C by the University of Brescia (UniBS). Upon obtaining participants' consent and following the co-signing of a Material Transfer Agreement and a Data Use Agreement compliant with the European General Data Protection Regulation by both institutions and the approval of a new IRB, biological samples and datasets were shared with the Department of Environmental health Sciences at the FIU Stempel College of Public Health and Social Work in 2023. Sample preparation and untargeted metabolomics and lipidomics methodology was described previously [25-29]. Briefly, whole blood samples were thawed at 4°C, vortexed for 5 seconds, and 50 µL of blood aliquot was transferred to a 1.5 mL Eppendorf tube. To this tube, 550 µL of methanol (MeOH) containing 200 µM 13C3-Lactate and 50 µM 13C5-15N-Glutamic Acid were added as the internal standards (ISs). The mixture was vortexed for 5 seconds and stored at -20°C for 20 minutes. It was then centrifuged at 14,000 rpm for 10 minutes, and 450 µL of the supernatant was collected. This sample was dried using an Eppendorf Vacufuge drier for 120 minutes, reconstituted in 150 µL of H2O:PBS:ACN (2:2:6) solution, and centrifuged again at 14,000 rpm for 10 minutes. A volume of 100 µL of the supernatant was transferred to a glass vial for metabolomics analysis. The remaining supernatant from each sample was allocated to a new tube to serve as a pooled quality control (QC) sample.

For lipidomics, starting with 50 µL of blood, 50 µL of 10X diluted PBS and 40 µL of 20X diluted Splash (Internal Standard Mixture) in MeOH were added. Then, 200 µL of Methyl tert-butyl ether (MTBE) was introduced to each sample in a ratio of MTBE/MeOH/H2O (10:2:5, v/v/v). The mixture was vortexed for 30 seconds and incubated at -20°C for 20 minutes, followed by centrifugation at 14,000 rpm for 10 minutes to induce phase separation. The upper MTBE layer (150 µL) was carefully pipetted into a new Eppendorf tube. The tubes were left open in a hood for 2 hours to dry. Afterward, 200 µL of Isopropanol/MeOH (1:1) was added to the dried residues, briefly sonicated, and centrifuged at 14,000 rpm for 10 minutes. A volume of 150 µL from each sample was then gently transferred to a glass vial for lipidomics analysis.

**Untargeted Metabolomic and Lipidomic Data Acquisition**

Mass spectrometry experiments were conducted using a Thermo UPLC-Exploris 240 Orbitrap MS (Waltham, MA) as described previously [25-29]. Briefly, each sample underwent dual injections for analysis in both positive and negative ionization modes, with an injection volume set at 1 µL. For metabolomics, chromatographic separation was achieved on a Waters XBridge BEH Amide column (150 x 2.1 mm, 2.5 µm, Waters Corporation, Milford, MA), utilized for both modes of ionization. The system operated with a flow rate of 0.3 mL/min, while the auto-sampler was maintained at 4ºC, and the column oven temperature was regulated at 40°C. The chromatographic mobile phase consisted of Solvent A (0.1% formic acid in a 95% $H_2O$/5% acetonitrile (ACN) mixture) and Solvent B (0.1% formic acid in a 95% ACN/5% H2O mixture), starting with a 0.5 min elution at 90% B, followed by a decrease to 40% at t = 10.5 min, holding for 2 min before gradually returning to 90% to accommodate subsequent injections. Lipidomics analyses employed a Waters XSelect HSS T3 column in both ionization modes, with a consistent flow rate of 0.3 mL/min. The mobile phase for lipidomics was formulated with Solvent A (0.1% formic acid in a 60% $H_2O$/40% ACN mixture) and Solvent B (0.1% formic acid in a 90% isopropanol/10% ACN mixture), initiating with an isocratic elution of 50% B for 3 minutes, then progressively increasing to 100% B over 12 minutes, maintained for 10 minutes, before decreasing to 50% for subsequent sample preparations. For mass spectrometric analysis, an electrospray ionization (ESI) source was employed to capture untargeted metabolomics data within the 70-1000 m/z range and lipidomics data within the 200-2000 m/z range. Peak identification in MS spectra leveraged our collection of approximately 300 in-house aqueous metabolite standards and involved searches across databases such as HMDB, mzCloud, Metabolika, and ChemSpider. Data extraction from MS required a minimum absolute intensity threshold of 1,000 and a mass accuracy within 5 ppm. Identifications and annotations were based on retention time, exact mass, MS/MS fragmentation, and isotopic patterns. Aqueous metabolomics data processing utilized Thermo Compound Discoverer 3.3 software for peak identification, alignment, and normalization, ensuring that only signals with a coefficient of variation (CV) less than 20% across quality control (QC) pools and those present in more than 80% of samples were analyzed further. Thermo LipidSearch 4.2 software was used for untargeted lipidomics data processing.

**Statistical Analysis**

*Sociodemographic and Covariate Data Analysis*

We report descriptive statistics for all covariates of the models, which were selected for their biological role on the exposure or the disease. The covariates include age (continuous), sex, coffee consumption (yes/no), alcohol consumption (yes/no), smoking status (ever/never), and comorbidities (none/at least one). Comorbidities were defined as the presence of any of the following conditions: diabetes, stroke, hypertension, leukemia, heart disease, liver disease, kidney disease, and thyroid disease. We performed Chi-square tests for categorical variables and t-tests for continuous variables to compare groups regarding disease status and exposure status. We also performed Spearman's rank correlation analysis to evaluate the monotonic relationships between exposure status, disease status, lifestyle choices, and demographic factors. Variables were ordinally ranked based on their contribution to disease risk. The rankings were informed by epidemiological studies, clinical research, and expert consensus. This non-parametric method was employed to determine correlation coefficients, reflecting the strength and direction of associations across the range of variables. All results were considered statistically significant with a Type I error of 5%.

*Metabolomic Data Analysis*

We used a 2x2 contingency table to investigate the metabolomic biomarker associations with 1) exposures (yes/no), 2) diagnosis of PD (yes/no) and 3) their interactions. We applied a series of computational techniques to analyze and interpret the data derived from 550 identified metabolites. To address the skewness of metabolite concentration data and to meet the model assumptions, a log10-transformation was implemented. Next, we performed outlier identification utilizing unsupervised (Principal Component Analysis [PCA]) and supervised dimensionality reduction (Partial Least Squares - Discriminant Analysis [PLS-DA]). To statistically evaluate the PCA results, a Permutational Multivariate Analysis of Variance (PERMANOVA) was conducted. For assessing the robustness of the PLS-DA model, 5-fold cross-validation metrics were calculated, permutation testing was performed, and Variable Importance in Projection (VIP) scores for each metabolite were analyzed. Metabolites with higher VIP scores were considered more influential in differentiating between groups. We further employed two-way Analysis of Covariance (ANCOVA) to discern the metabolomic alterations of 1) exposures (yes/no), 2) diagnosis of PD (yes/no) and 3) their interactions while adjusting for covariates (age and sex). Raw and adjusted P-values using the False Discovery Rate (FDR) Benjamini-Hochberg method were reported. Beta coefficients, 95% confidence intervals (CI), and partial Eta squared $n_p^2$) were also reported. $n_p^2$ value represents the proportion of total variance in metabolite levels that is attributable to each main effect or their interaction, after accounting for other variables included in the model. $n_p^2$ values were hierarchically ranked to ascertain a rank ordered list of metabolites. Statistical significance was determined using an 5% FDR threshold. To evaluate the biological relevance of our significant features, we performed overrepresentation pathway analysis (OPA) and metabolite set enrichment analyses (MSEA) using statistically significant metabolites as our test set. Organism-specific pathway sets were used as our background set for pathway analysis (Homo Sapien - KEGG) and enrichment analysis (Homo Sapien - RaMP-DB) as suggested by previous reports [30, 31]. Using a hypergeometric test for enrichment and relative-betweenness centrality for topology analysis, pathways were identified based on statistical significance and impact score (Impact) associated with each main effect or the interaction between these factors. Pathways with an uncorrected Type I error of 5% were considered biologically relevant. The enrichment analysis utilized the GlobalTest and topology analysis via the relative-betweenness-centrality method, which identified metabolites that were significantly enriched, considering their position within the metabolic network. The 'Expect' column represents the expected number of metabolite hits in each set based on a random distribution, calculated under the null hypothesis. This value provides a baseline for comparing the observed number of hits, allowing for the assessment of whether the metabolite set is significantly enriched beyond what would be expected by chance." All statistical analyses and visualizations were performed using a combination of software tools including RStudio (Version 2023.09.1+494), MetaboAnalyst (Version 6), and Jupyter Notebooks (Version 6.5.4) accessed through Anaconda.

*Lipidomic Data Analysis*

We applied a series of computational techniques to analyze and interpret the data derived from 94 identified lipid species following the analysis methodology described above for metabolomic data. Note, biological relevance was assessed using an alternative software tool, LIPEA. Briefly, LIPEA leverages the KEGG Database to perform OPA. OPA was conducted using the Fisher's exact test to identify statistically significant lipids. These lipids showed overrepresentation when compared to the KEGG Database list of lipids, which served as the background dataset. The 'Pathway Lipids' column represents the number of lipids associated with each specific pathway that were tested for enrichment. To determine whether these pathways are significantly enriched with the lipids identified in our dataset, we applied Fisher's Exact Test. This test assesses the association between the identified lipids and the pathways, comparing the observed number of lipids within each pathway to what would be expected under a null hypothesis of no association. This background set included all lipids that could potentially be identified in the specific organism under study, Homo sapiens. Biologically relevant overrepresented pathways were identified using the 5% FDR threshold. Note, due to a lack of statistically significant features identified for the exposure effect, biological relevance was not assessed using OPA. All statistical analyses and visualizations were performed using a combination of software tools including RStudio (Version 2023.09.1+494), LIPEA (https://hyperlipea.org/home), and Jupyter Notebooks (Version 6.5.4) accessed through Anaconda.

Supplemental Figure 1: Correlation Matrix and P-Value Matrix for Selected Variables

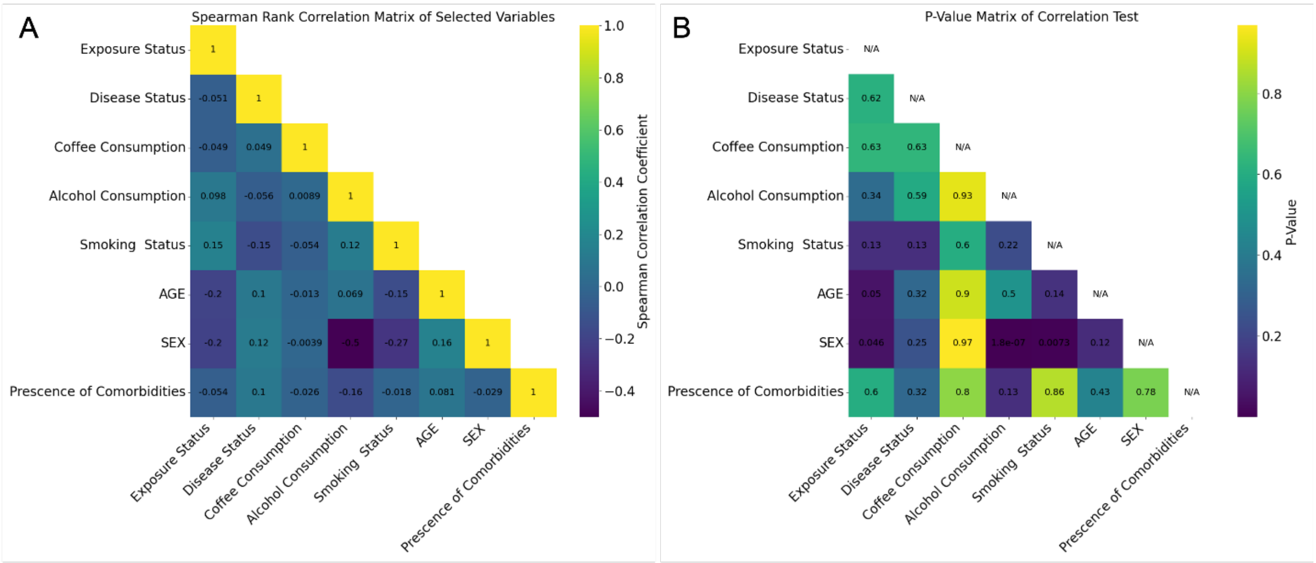


Panel A: Spearman rank correlation matrix of selected variables, showing the correlation coefficients between exposure status, disease status, coffee consumption, alcohol consumption, smoking status, age, sex, and the presence of comorbidities. The color gradient represents the strength and direction of correlations. Panel B: P-value matrix of the Spearman rank correlation test for selected variables. The color gradient indicates the significance levels of the correlations, with non-significant (n.s.) values marked accordingly. The strongest positive correlation was found between Sex and Age (0.16), although this correlation was weak and not statistically significant ($p < 0.05$). The strongest negative correlations were found between sex and alcohol consumption (-0.50) and sex and smoking status (-0.27). Upon further investigation, sex contrast against alcohol consumption and sex contrast against smoking status show statistically significant correlations ($p < 0.05$), with males being more likely to smoke and consume alcohol compared to females in this dataset. Most notably, age and sex show a negative correlation with exposure (-0.2 and -0.2, respectively) and borderline statistical significance ($p = 0.04$ and $p = 0.05$, respectively) indicating that age and sex covary with exposure. From these heatmaps, most lifestyle and demographic factors do not have a strong or significant correlation with exposure status or disease status (our outcomes of interest), apart from age and sex, which have marginal levels of significant association with exposure. Given these findings we included covariate adjustments for age and sex in all subsequent analyses.

Supplementary Figure 2: Lifestyle Habits by Sex

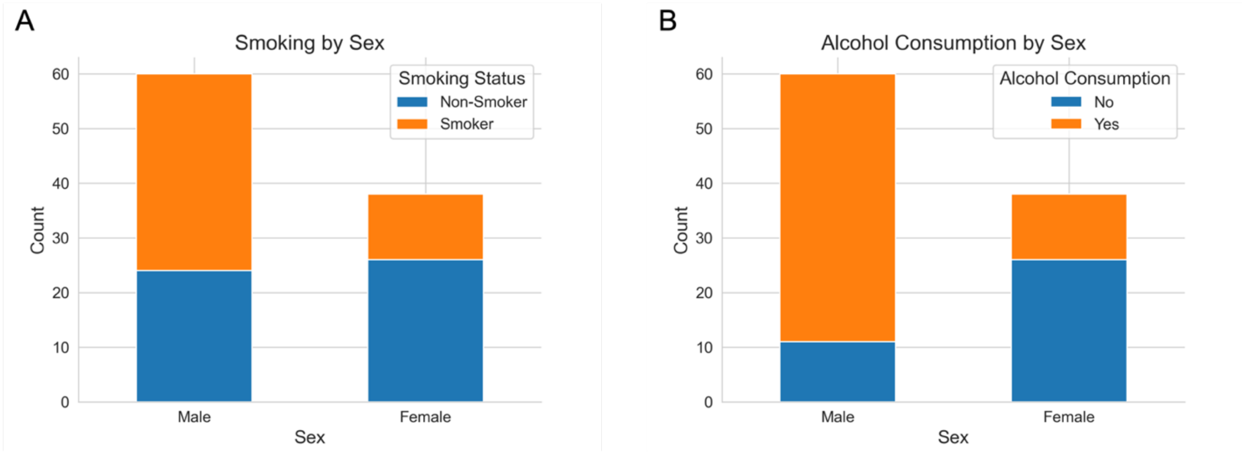


Panel A: Smoking status by sex, showing counts of smokers and non-smokers among male and female participants. Panel B: Alcohol consumption by sex, displaying the number of participants who consume alcohol versus those who do not, categorized by sex.

Supplemental Figure 3: VIP Scores and Relative Abundance of Significant Metabolites

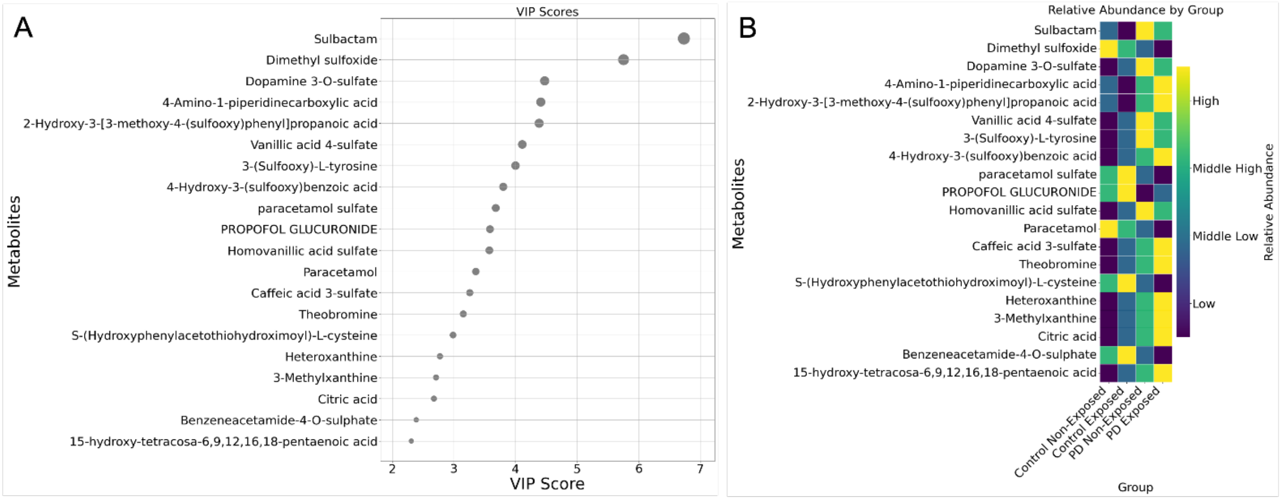


Panel A: VIP scores for the top 20 significant metabolites, showing their importance in differentiating between the groups. Higher VIP scores indicate greater importance in the model. Panel B: Heatmap of relative abundance of the top 20 significant metabolites by group. The color gradient represents the abundance levels, with darker colors indicating higher abundance. The groups compared include control non-exposed, control exposed, PD non-exposed, and PD exposed. PLS-DA generated a ranked-ordered list of metabolites based on their Variable Importance in Projection (VIP) scores (*Panel A*) and associated relative abundance (*Panel B*). The metabolite with the most substantial influence on the model was sulbactam, registering a VIP score above 6. This was followed by “dimethyl sulfoxide” and “dopamine 3-O-sulfate”, each with VIP scores ranging between 4 and 6. All other metabolites in the top 20 list presented VIP scores above 2, with “15-hydroxy-tetracosa-6,9,12,16,18-pentaenoic acid” concluding the list.

Supplemental Table-Figure 1: PLS-DA Cross Validation Metabolites

**PLS-DA cross validation**

| Measure | 1 comps | 2 comps | 3 comps | 4 comps | 5 comps |
|---|---|---|---|---|---|
| Accuracy | 0.44368 | 0.56737 | 0.69 | 0.72105 | 0.75158 |
| R2 | 0.57214 | 0.80643 | 0.89846 | 0.9423 | 0.9605 |
| Q2 | 0.47213 | 0.60772 | 0.68003 | 0.74047 | 0.75471 |

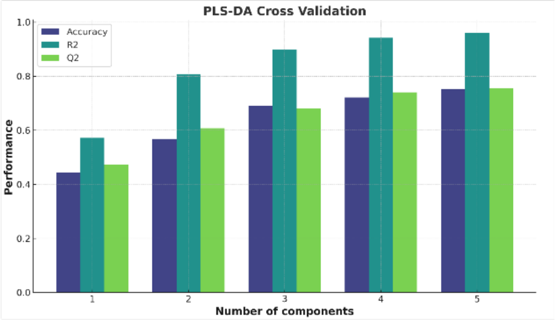


Table: Table showing PLS-DA cross-validation metrics, including accuracy, R2, and Q2 values for models with 1 to 5 components. Figure: Bar chart illustrating PLS-DA cross-validation results for accuracy, R2, and Q2 across different numbers of components. The number of components is represented on the x-axis, and the performance measures are represented on the y-axis, with different colors indicating the specific measures. To evaluate the model's performance, we utilized 5-fold cross-validation with varying numbers of components, along with permutation testing. Permutation testing with 1000 permutations using prediction accuracy during training as the test statistic resulted in a p-value less than 0.001, indicating that the model's performance is significantly better than what would be expected by random chance (Data Not Shown). In addition to prediction accuracy, we evaluated the separation distance (B/W) to understand the structural differences between the classes in the latent space. The separation distance provided further insights into the discriminative power of the model, complementing the accuracy metrics. The resulting p-value was also less than 0.001, suggesting a statistically significant separation (Data Not Shown).

Supplementary Figure 4: Distribution of Metabolite Concentrations

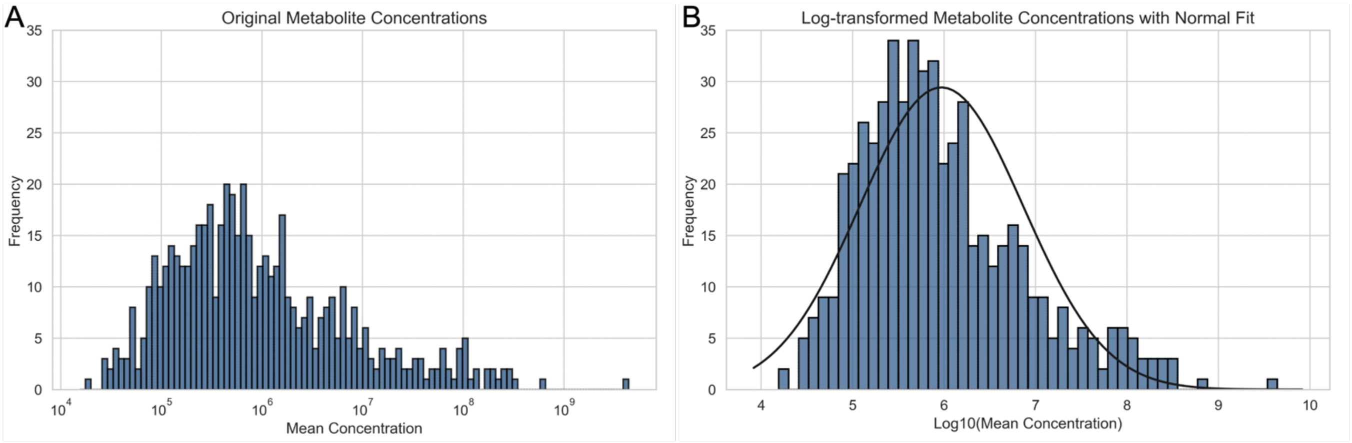


Panel A: Histogram of original metabolite concentrations, displaying the frequency distribution of metabolite levels before transformation. Panel B: Histogram of log-transformed metabolite concentrations with normal fit, showing the distribution of metabolite levels after log transformation and overlaid with a normal distribution curve for comparison.

Supplementary Figure 5: Scores Plots for PCA and PLS-DA Analysis Metabolites

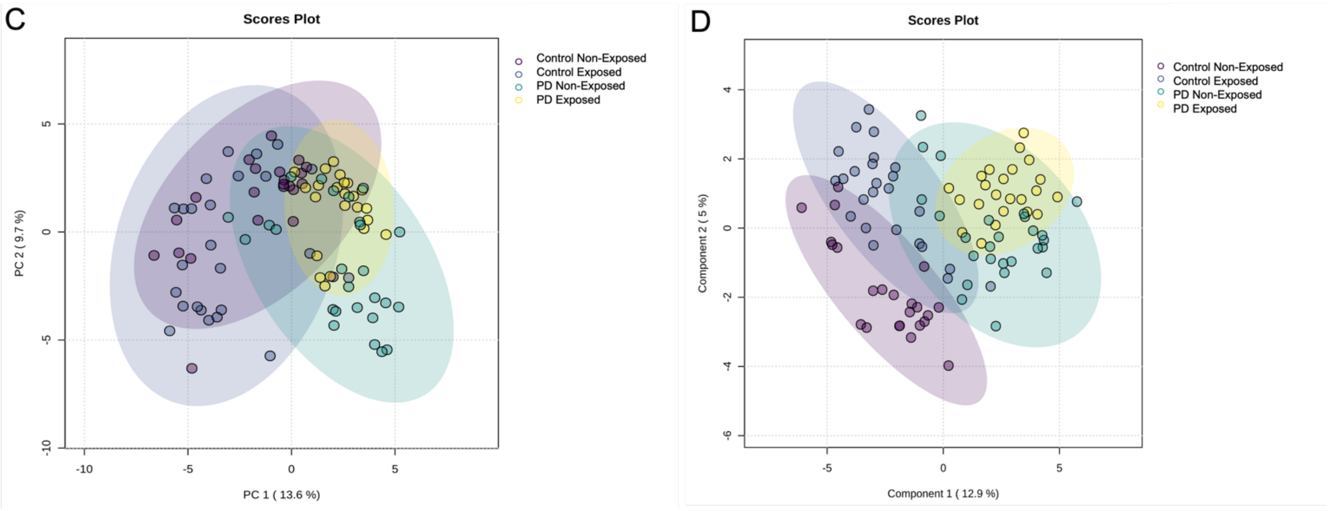


Panel C: PCA scores plot showing the first two principal components (PC1 and PC2) for control and Parkinsonism (PD) subjects, both exposed and non-exposed. PC1 and PC2 capture the largest variance in the dataset, with PC1 accounting for 13.6% and PC2 for 9.7%. PERMANOVA analysis was conducted, yielding an F-value of 22.665, an R-squared value of 0.42234, and a p-value of 0.001 (based on 999 permutations) (Data Not Shown). These PERMANOVA results indicate a strong and statistically significant differentiation between groups. Panel D: PLS-DA scores plot also displays the first two components (PC1 and PC2) for control and PD subjects, highlighting more distinct group separations compared to PCA with PC1 explaining 12.9% and PC2 explaining 5% of the variance. PLS-DA, being a supervised technique, shows a clearer separation between the groups, particularly between the Control and PD groups along Component 1. This indicates that the model can use the group labels to find metabolites that differentiate between disease and non-disease states. The PD Exposed group is more distinct from the Control Non-Exposed group, suggesting that exposure status may play a role in the metabolomic distinction of PD. The overlap between the Control Exposed and PD Non-Exposed groups suggests that exposure might have a metabolomic signature that is like the signature of PD without exposure.

Supplementary Figure 6: Venn Diagrams of Significant Metabolites

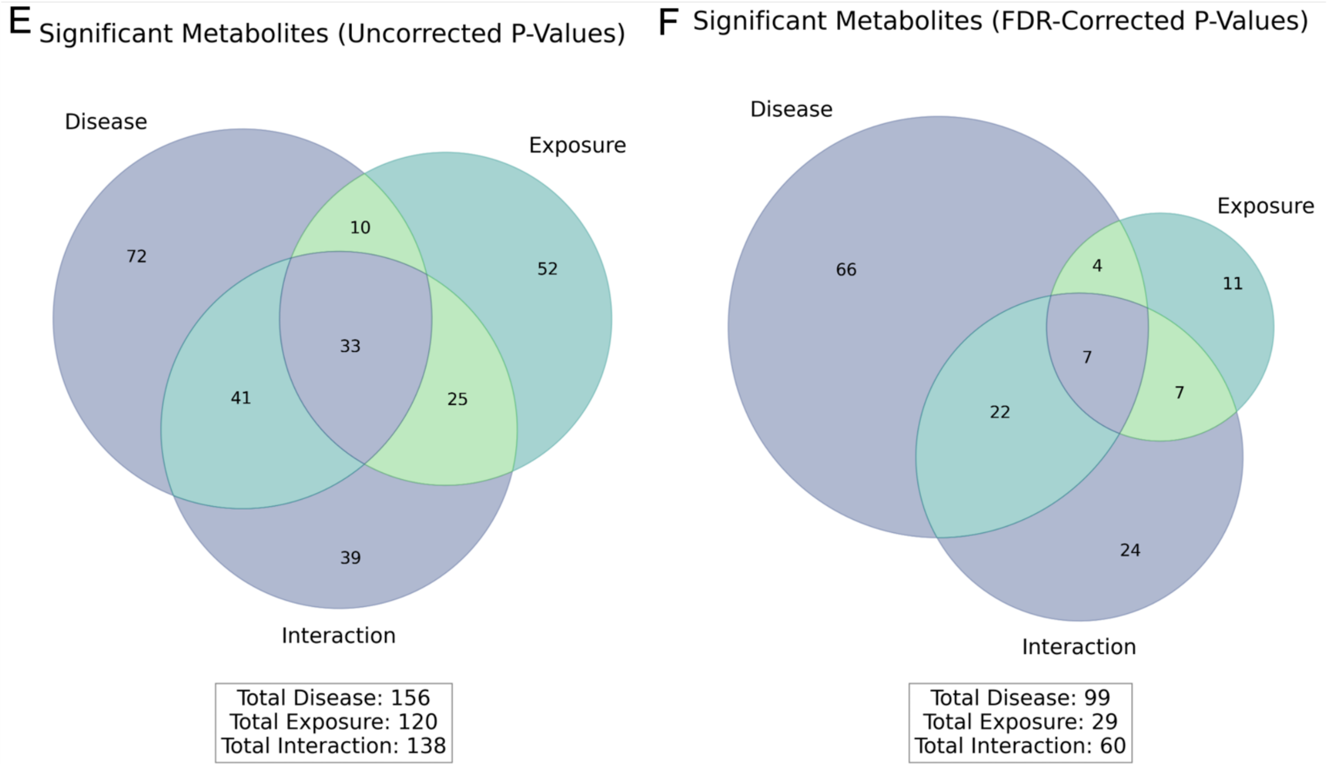


Panel E: Venn diagram showing the overlap of significant metabolites for disease, exposure, and interaction effects based on uncorrected p-values. Total significant metabolites are listed for each category. Panel F: Venn diagram displaying the overlap of significant metabolites for disease, exposure, and interaction effects based on FDR-corrected p-values. Total significant metabolites are listed for each category after FDR correction.

Figure 7: VIP Scores and Relative Abundance of Significant Lipids

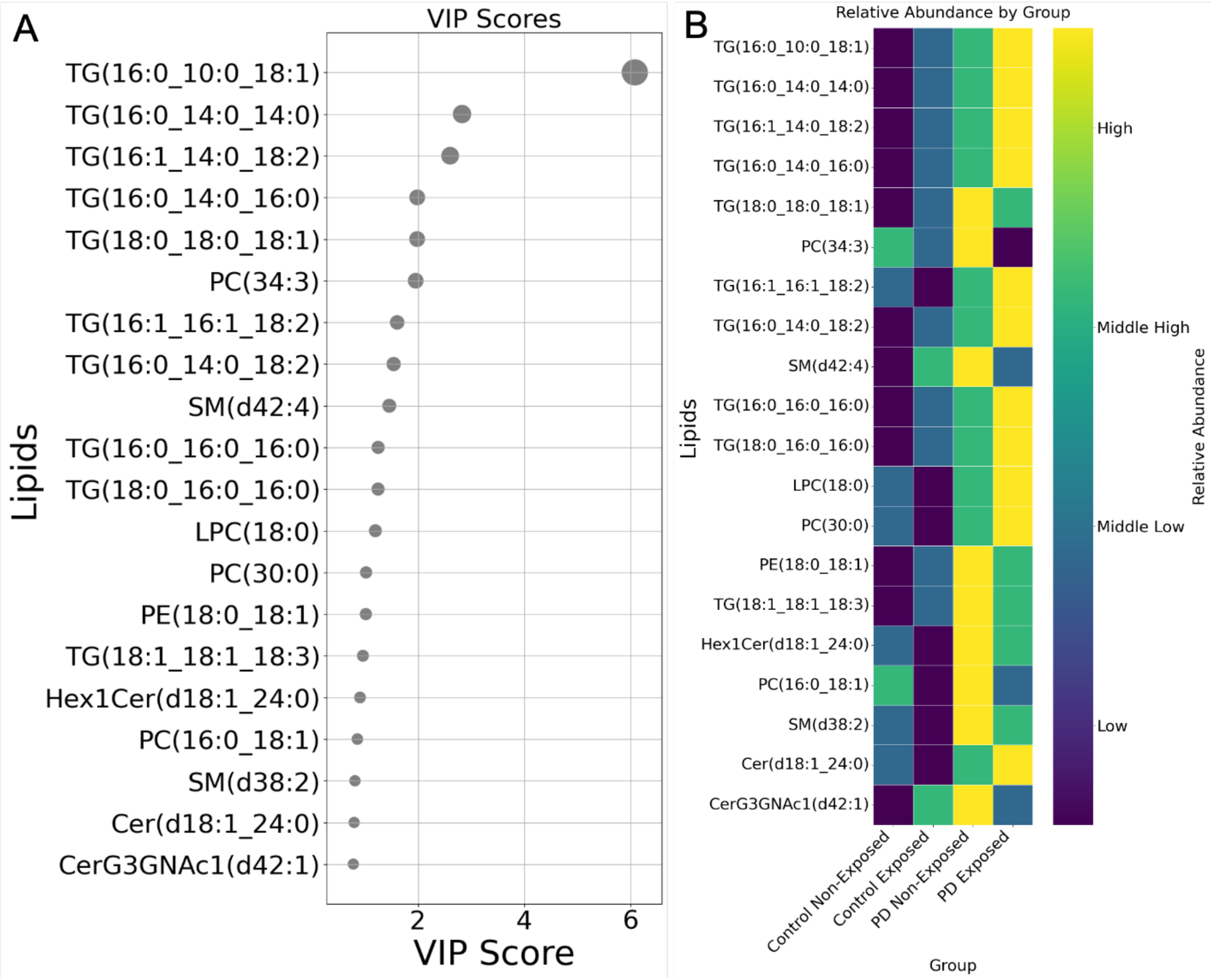


Panel A: VIP scores for the top 20 significant lipids, showing their importance in differentiating between the groups. Higher VIP scores indicate greater importance in the model. Panel B: Heatmap of relative abundance of the top 20 significant lipids by group. The color gradient represents the abundance levels, with darker colors indicating higher abundance. The groups compared include control non-exposed, control exposed, PD non-exposed, and PD exposed. PLS-DA generated a ranked ordered list of lipids based on their Variable Importance in Projection (VIP) scores (*Panel A*) and associated relative abundance (*Panel B*). “TG(16:0_10:0_18:1)”, “TG(16:0_14:0_14:0)”, and “TG(16:1_14:0_18:2)” are the top three lipids based on VIP scores (6, ~3, and ~2.5, respectively), highlighting their significant roles in discriminating between the groups. The relative abundance heatmap shows a clear trend of increasing levels of these lipids from Control Non-Exposed to PD Exposed, suggesting their strong association with PD status and exposure.

Supplemental Table-Figure 2: PLS-DA Cross Validation Lipids

### PLS-DA cross validation

| Measure | 1 comps | 2 comps | 3 comps | 4 comps | 5 comps |
|---|---|---|---|---|---|
| Accuracy | 0.38779 | 0.51418 | 0.54576 | 0.57687 | 0.54423 |
| R2 | 0.38774 | 0.50138 | 0.52724 | 0.57203 | 0.61322 |
| Q2 | 0.33009 | 0.24817 | 0.27351 | 0.25271 | 0.14685 |

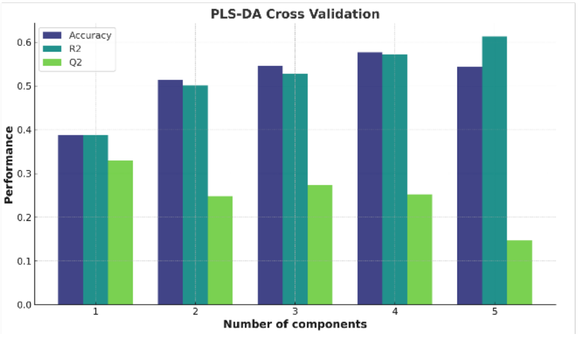


Table: Table showing PLS-DA cross-validation metrics, including accuracy, R2, and Q2 values for models with 1 to 5 components. Figure: Bar chart illustrating PLS-DA cross-validation results for accuracy, R2, and Q2 across different numbers of components. The number of components is represented on the x-axis, and the performance measures are represented on the y-axis, with different colors indicating the specific measures. To evaluate the model's performance, we utilized 5-fold cross-validation with varying numbers of components, along with permutation testing. Permutation testing with 1000 permutations using prediction accuracy during training as the test statistic resulted in a p-value equal to 0.082, indicating that the model's performance is not significantly better than what would be expected using a Type I error of 5% (Data Not Shown). In addition to prediction accuracy, we evaluated the separation distance (B/W) to understand the structural differences between the classes in the latent space. The separation distance provided further insights into the discriminative power of the model, complementing the accuracy metrics. The resulting p-value was less than 0.001, suggesting statistically significant class separation (Data Not Shown). These lipids may serve as potential biomarkers for PD, especially in individuals with exposure, and warrant further investigation for their biological roles and mechanism.

Supplementary Figure 8: Pathway Analysis and Metabolite Set Enrichment Overview

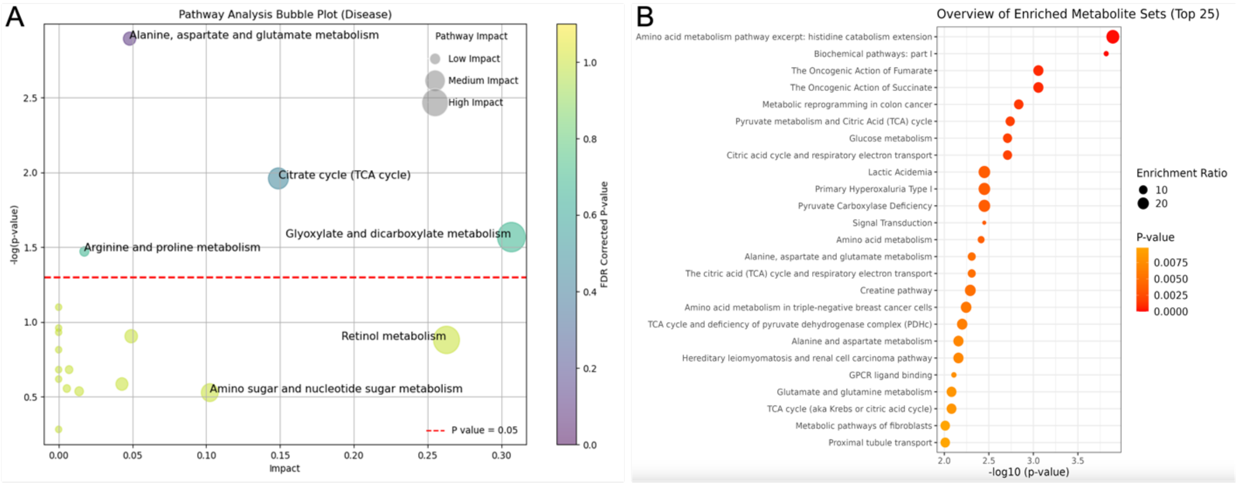


Panel A: Pathway analysis bubble plot for disease effect, showing the significance and impact of various metabolic pathways. Pathways with high impact and significance include "Alanine, aspartate and glutamate metabolism" and "Glyoxylate and dicarboxylate metabolism." The color gradient represents the FDR corrected p-value, with pathways above the red dashed line (p-value = 0.05) considered statistically significant. Panel B: Overview of the top 25 enriched metabolite sets. The size of the bubbles represents the enrichment ratio, and the color gradient indicates the p-value. More significant pathways are highlighted with larger and darker-colored bubbles.

Supplementary Figure 9: Top Metabolites and Enriched Metabolite Sets for Exposure Effect

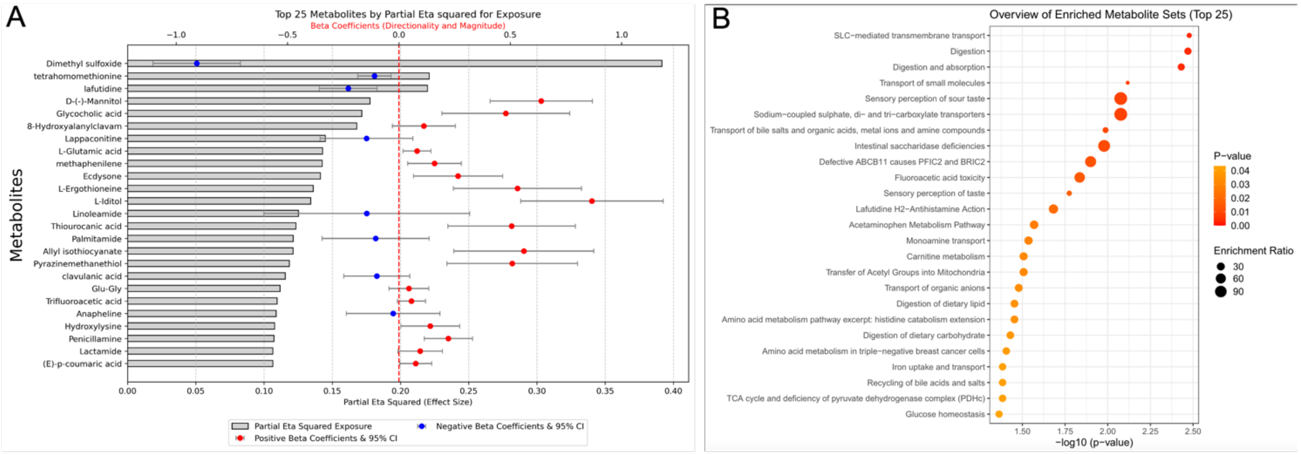


Panel A: The top 25 metabolites by partial eta squared for the exposure effect. The bar plot displays partial eta squared values (effect sizes), with positive and negative beta coefficients indicated by red and blue dots, respectively, along with their 95% confidence intervals. Panel B: Overview of the top 25 enriched metabolite sets. The bubble plot represents the enrichment ratio and significance of various metabolite sets, with the size of the bubbles indicating the enrichment ratio and the color gradient representing the p-value. More significant pathways are highlighted with larger and darker-colored bubbles.

Supplementary Figure 10: Top Metabolites and Pathways for Interaction Effect

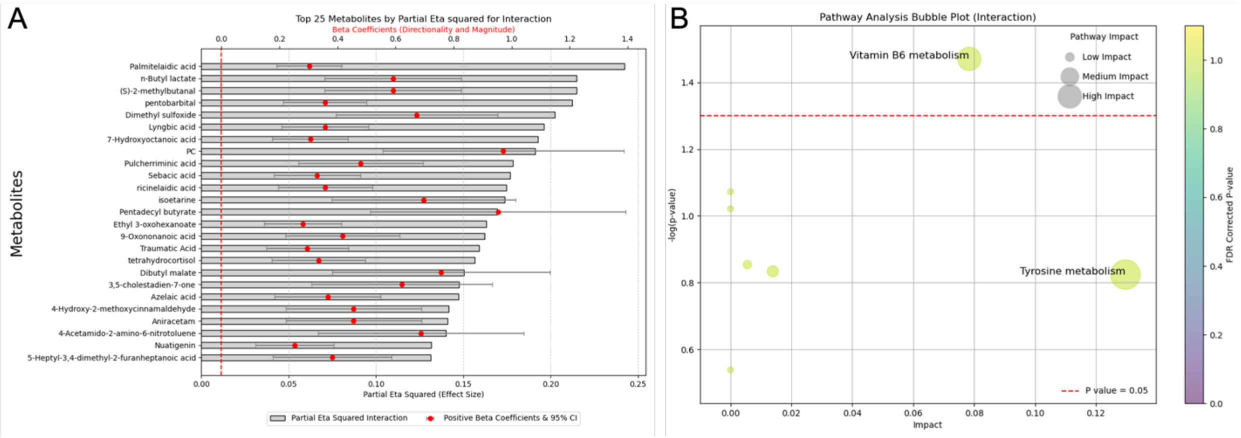


Panel A: Top 25 metabolites by partial eta squared for the interaction effect. The bar plot displays partial eta squared values (effect sizes), with positive beta coefficients indicated by red dots along with their 95% confidence intervals. Panel B: Pathway analysis bubble plot for the interaction effect. The bubble plot shows the impact and significance of pathways affected by the interaction effect, with bubble size representing pathway impact and color gradient indicating FDR-corrected p-values. The red dashed line represents the p-value threshold of 0.05.

Supplemental Figure 11: Distribution of Lipid Concentrations

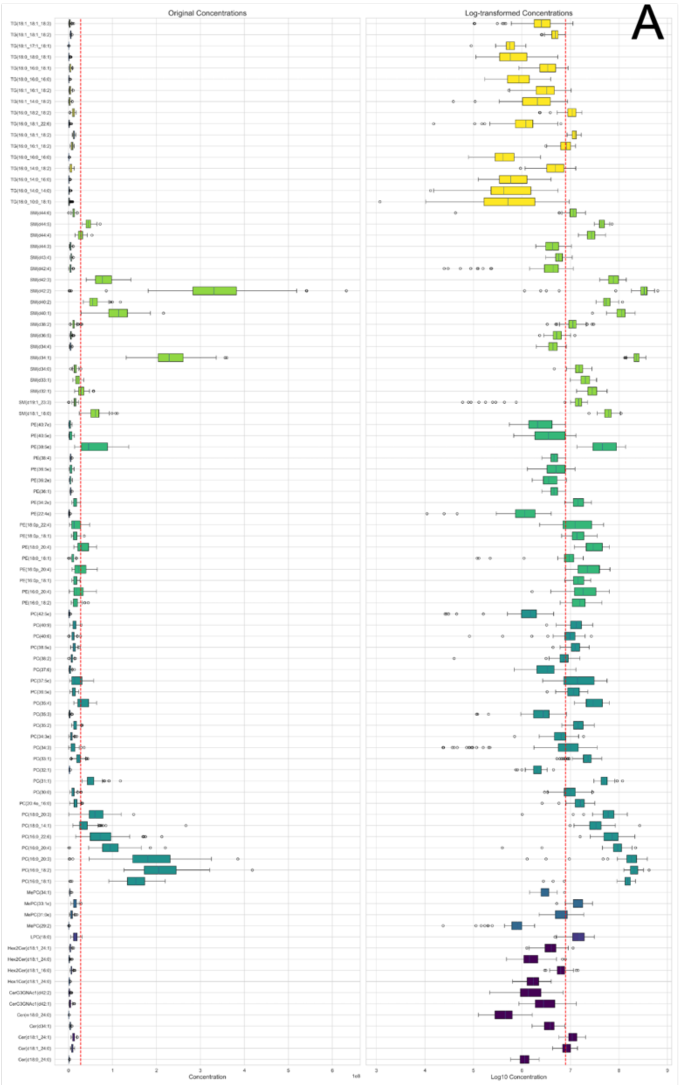


Left Panel: Original observations of lipid concentrations. The box plot displays the spread and skewness of lipid concentration data before log-transformation, with individual outliers marked. Right Panel: Log-transformed lipid concentrations. The box plot shows the same lipid concentration data after log-transformation, illustrating the normalization effect. The red dashed line represents the median value for comparison between original and log-transformed data.

Supplemental Figure 12: Scores Plots for PCA and PLS-DA Analysis Lipids

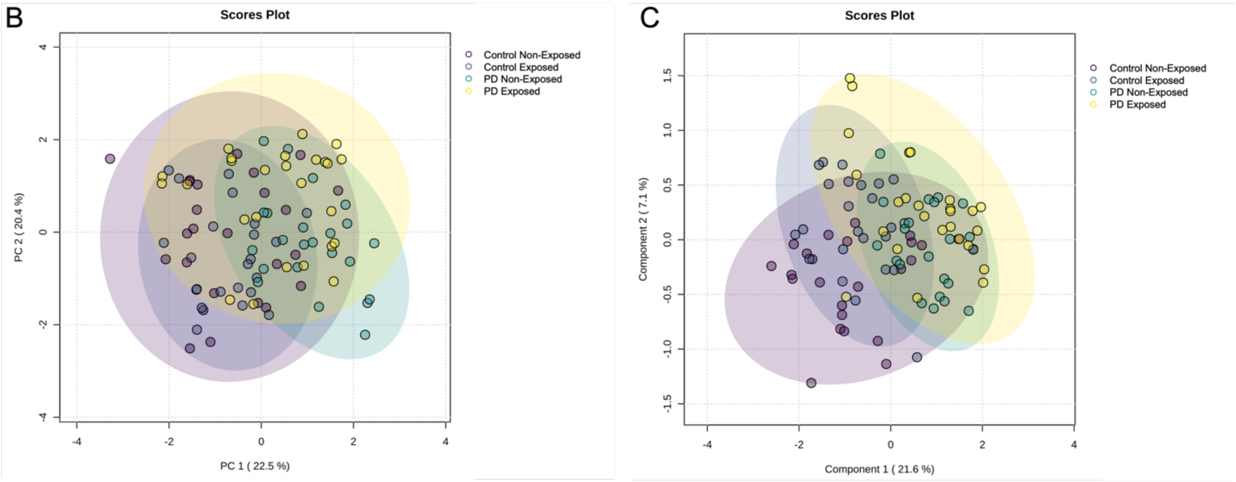


Panel B: Principal Component Analysis (PCA) plot displaying the first two principal components (PC1 and PC2) for control and PD subjects, both exposed and non-exposed. PC1 accounts for 22.5% and PC2 accounts for 20.4% of the variance. Each point represents a subject, colored by group, with 95% confidence ellipses. PERMANOVA analysis was conducted, yielding an F-value of 10.922, an R-squared value of 0.26053, and a p-value of 0.001 (based on 999 permutations) (Data Not Shown). These PERMANOVA results indicate a strong and statistically significant differentiation between groups. Panel C: Partial Least Squares Discriminant Analysis (PLS-DA) plot displaying the first two components for control and PD subjects, both exposed and non-exposed. Component 1 accounts for 21.6% and Component 2 accounts for 7.1% of the variance. Each point represents a subject, colored by group, with 95% confidence ellipses.

Supplemental Figure 13: Venn Diagrams of Significant Lipids

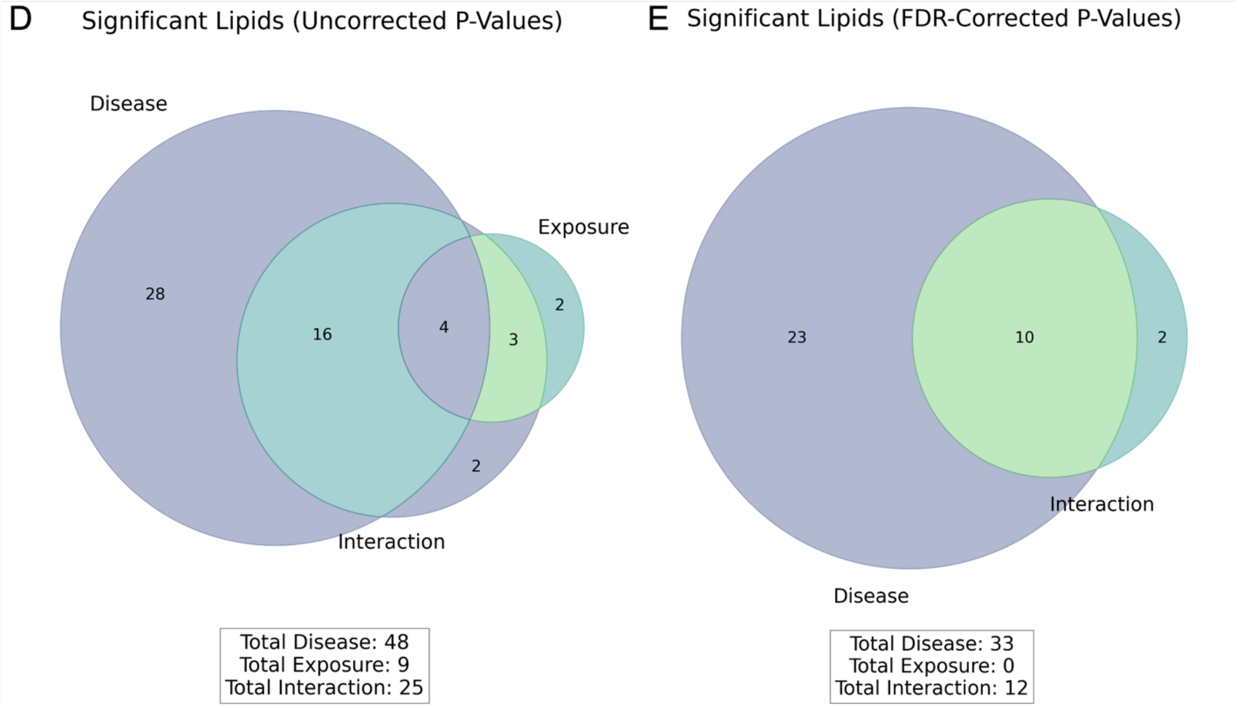


Panel D: Venn diagram showing the number of significant lipids associated with disease, exposure, and interaction effects before False Discovery Rate (FDR) correction. The overlap represents lipids significant across multiple effects. Total significant lipids are listed below the diagram: Disease (48), Exposure (9), and Interaction (25). Panel E: Venn diagram showing the number of significant lipids associated with disease, exposure, and interaction effects after FDR correction. The overlap represents lipids significant across multiple effects. Total significant lipids are listed below the diagram: Disease (33), Exposure (0), and Interaction (12).
